# Neocortical Localization and Thalamocortical Modulation of Neuronal Hyperexcitability in Fragile X Syndrome

**DOI:** 10.1101/2021.05.12.21256925

**Authors:** Ernest V Pedapati, Lauren M. Schmitt, Lauren E. Ethridge, Rui Liu, Elizabeth Smith, John A Sweeney, Rebecca C. Shaffer, Kelli C. Dominick, Donald L. Gilbert, Steve W. Wu, Paul S. Horn, Devin Binder, Martine Lamy, Megan Axford, Makoto Miyakoshi, Craig A. Erickson

## Abstract

Fragile X Syndrome (FXS) is a monogenetic form of intellectual disability and autism in which well-established knockout (KO) animal models point to neuronal hyperexcitability and abnormal gamma-frequency physiology as a basis for key disorder features. Translating these findings into patients may identify tractable treatment targets. Using source modeling of resting-state electroencephalography data, we report novel findings in FXS, including 1) increases in localized gamma activity, 2) pervasive changes of theta/alpha activity, indicative of disrupted thalamocortical modulation coupled with elevated gamma power, 3) stepwise moderation of these abnormalities based on female sex, and 4) relationship of this physiology to intellectual disability and neuropsychiatric symptoms. Our observations extend findings in *Fmr1*^-/-^ KO mice to patients with FXS and raise a novel role for disrupted thalamocortical modulation in local hyperexcitability. This systems-level mechanism has received limited preclinical attention but has significant implications for understanding fundamental disease mechanisms.

## Introduction

Loss of the fragile X mental retardation protein (FMRP), mainly caused by the silencing of the fragile X mental retardation 1 (*Fmr1*) gene, leads to Fragile X Syndrome (FXS)^1^. FXS is an X-linked disorder that results in near-universal intellectual disability with a high prevalence of anxiety disorders, communication impairments, sensory hypersensitivities, and autism. FMRP is a polyribosome-associated RNA binding protein that regulates the levels of many pre- and postsynaptic proteins^2^. FMRP indirectly regulates proteins that maintain neuronal excitability and directly interact with membrane-bound ion channels^3^. The loss of FMRP has been associated with neuronal hyperexcitability. The *Fmr1*^-/-^ knockout (KO) mouse, for example, is susceptible to audiogenic seizures, displays increased spontaneous network spiking, and elevation in resting-state electroencephalography (EEG) gamma power (> 30 Hz)^4-7^. As the *Fmr1*^-/-^ KO is central to preclinical development, it is essential to understand parallels in humans with the mouse model, but there is also an urgent need to extend our understanding of system-level dynamics in humans,

EEG is a highly feasible method to postulate whole-brain hypotheses in populations such as FXS, where invasive recordings are not available, and other neuroimaging may pose selection bias towards less impaired participants^8^. Previous reports of EEG activity in FXS have consistently demonstrated abnormalities in the alpha, theta, and gamma band. Changes in low-frequency power have been reported from the earliest clinical EEG studies of FXS, including elevated theta power and reduced alpha power^9-12^. Additionally, as in the *Fmr1*^-/-^ KO, humans with FXS manifest increased gamma power^12-14^. Such low and high-frequency changes may also be linked, as individuals with FXS display an increase in theta-gamma cross-frequency power-power coupling (CFC) during resting state EEG^12^. Despite these intriguing findings, the confirmation of system-level hypotheses and clinical correlations have been limited by small samples (n<25), underrepresentation of females, failure to ascertain mosaic status in males, and lack of source and functional network modeling.

Changes in thalamocortical activity are a system-level hypothesis that could explain these EEG findings and, thus far, underexplored in FXS and preclinical models. Thalamocortical dysrhythmia (TCD) is an electrophysiological motif derived from magnetoencephalography and EEG that has been attributed to the dysregulation of cortical excitability and observed across neuropsychiatric conditions (i.e., epilepsy, Parkinson’s disease, tinnitus, depression, and neuropathic pain)^15-17^. TCD related EEG alterations have been associated with clinical features in these disorders, and most recently in the schizophrenia^18^. Like in FXS, patient groups in TCD display have reduced alpha power, increased theta power, increased gamma power, and predominance of theta gamma CFC.

Herein, we provide evidence of TCD in FXS. In comparison to previous reports^14^, we have substantially increased the sample size, conducted source localization, and modeled cortical regions and functional networks. We expected to confirm changes in theta, alpha, and gamma activity signatures consistent with TCD. Finally, we expected to find evidence that TCD-related alterations would demonstrate clinical correlation with core disorder features, including cognition and neuropsychiatric symptoms. These findings provide parallels to the *Fmr1*^-/-^ KO and implicate subcortical contributions to the pathophysiology of FXS, which has thus far been under-reported in the literature.

## Results

### Participants

We compared eighty seconds of artifact-free resting-state EEG data (**see Figure 1**) between 70 individuals with a genetic diagnosis of FXS (without seizures or on antiepileptics) and 71 age- and sex-matched typically developing control participants (**see Table 1**). Other clinical phenotypic differences between groups were estimated by neuropsychiatric measures (**see Table S1**). Raw data were handled blindly, and there were no differences in preprocessing characteristics between groups (**see Table S2**). To optimize the detection of neurogenic activity from the gamma band, we followed best practices to address myogenic contamination^19^. Unless otherwise specified, dependent variables that were analyzed by linear mixed effect model (LME) included group (FXS or control), sex (male or female), and frequency band (delta, theta, alpha1, alpha2, beta, gamma1, and gamma2) as fixed effects and subject as the random effect. We have organized the results to report three primary analyses: 1) changes in spectral power, 2) evaluation PAF, and 3) alterations of CFC. We conclude with a summary of clinical correlations across analyses.

**Figure 1:**
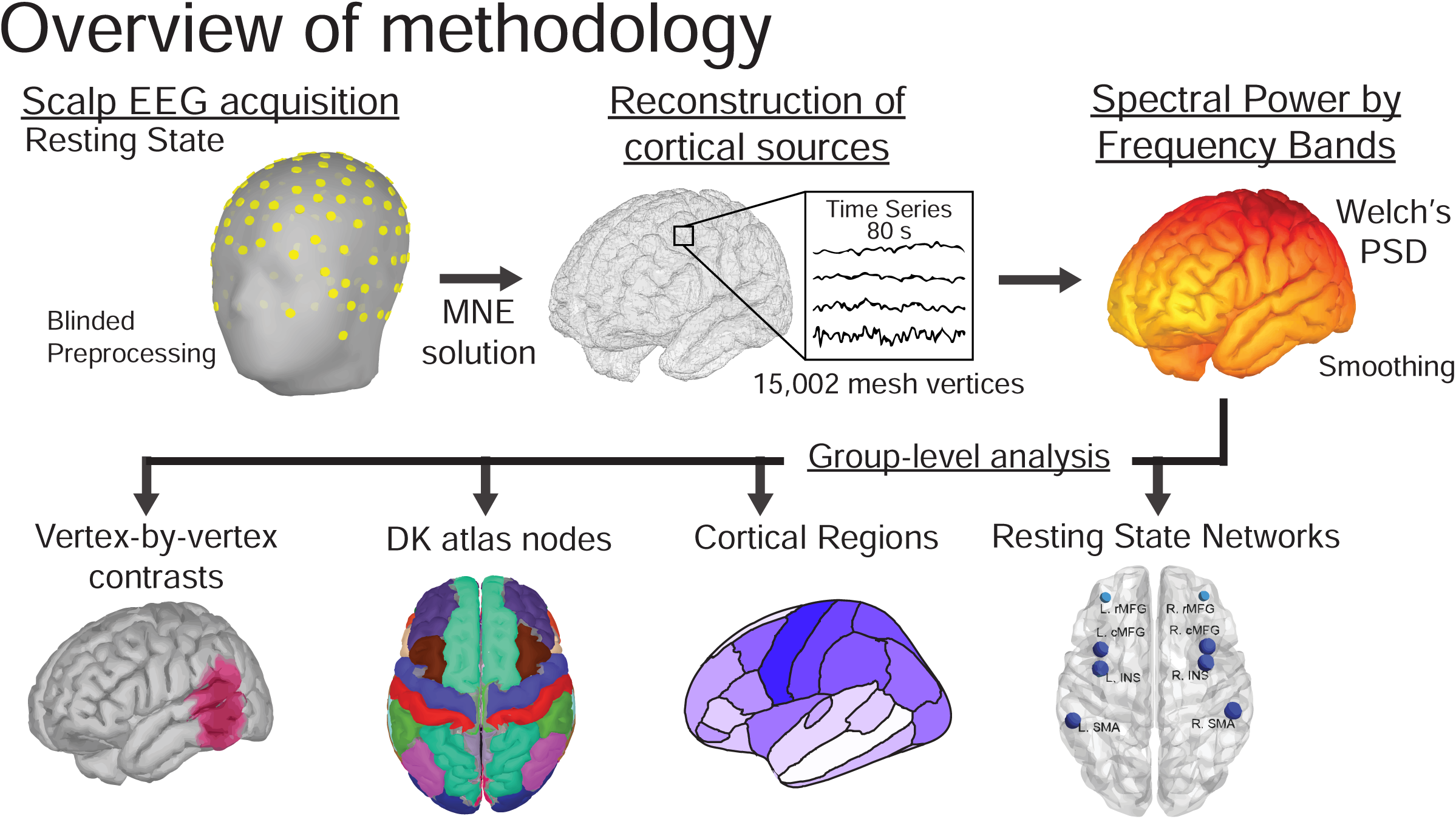
Principal steps of investigation: Following blinded preprocessing, artifact-free EEG data and a cortical lead field matrix was used to construct a weighted minimum norm estimate (MNE) source model to estimate current source density (CSD) and spectral power by frequency band at each vertex of a triangular cortical mesh (top row). Based on hypotheses, subsequent analyses were performed at four different levels (bottom row): raw vertices, parcellation of vertex points into Desikan-Killiany atlas-based nodes, cortical regions (anatomical continuous grouping of nodes), and/or resting-state networks (RSN) (functional non-continuous grouping of nodes).

**Table 1:** Demographic and clinical features of the EEG dataset showing mean (± standard deviation) group t-tests by group. FSIQ, Full Scale IQ, NVIQ, Non-verbal intelligence scale; VIQ, verbal intelligence scale; SCQ, Social Communication Questionnaire; WJ-3, Woodcock III Tests of Cognitive Abilities; ADAMS, Anxiety, Depression, and Mood Scale; t, t-statistic following independent Student t-tests; p, unadjusted significance; adj. p, p-value following Bonferroni correction.

| Measure | FXS (n=70) | Controls (n=71) | t | p | adj. p. |
| --- | --- | --- | --- | --- | --- |
| Age (Years) | 20.5±10.0 | 22.2±10.7 | <.001 | .35 | .70 |
| Sex (Female) | F: 32 M: 38 | F: 30 M: 41 | .17 | .72 | .72 |
| FSIQ | 49.5±30.2 | 103.2±9.2 | <.001 | <.001 | <.001 |
| NVIQ | 40 ± 35.7 | 103.4 ± 10.7 | -12.56 | <.001 | <.001 |
| VIQ | 58.9 ± 28.7 | 103.1 ± 12.8 | -10.45 | <.001 | <.001 |
| SCQ | 13.6±7.7 | 2.1±2.2 | 9.94 | <.001 | <.001 |
| WJ-III | 67.8±15.7 | 94.1±12.1 | <.001 | <.001 | <.001 |
| ADAMS-Anxiety | 7.2±5.0 | 2.1±2.6 | 6.37 | <.001 | <.001 |
| ADAMS-OCD | 2.3±2.5 | .4±1.1 | 5.12 | <.001 | <.001 |

### Evidence of reduced alpha, increased theta, and gamma power in FXS from scalp EEG

#### Scalp EEG Relative Power

Prior to source modeling, we considered the temporal and spatial properties of the scalp EEG recording by examining the topographical distribution and scalp-averaged spectrograms of relative power across groups. Relative power (proportion of band power to total power) is generally reported to reduce inter-subject variation and facilitate group comparisons^20^ in human studies. *Scalp topography:* We first examined power split by frequency bands between FXS and control groups. We found that in FXS, widespread alpha power decreases, theta power increases, and clusters of increased gamma power (**see Figure S1A**). *Scalp spectrogram and peak alpha frequency (PAF):* As in other examples of TCD, visual inspection revealed a global leftward shift in PAF towards the theta band in males and females with FXS (**see Figure 2B**). An LME confirmed that PAF was significantly reduced in FXS with a significant group x sex interaction effect (F_1,137_=5.597; p=.02), but no effect of electrode region. Estimates of PAF (mean Hz ± std. error) in male participants with FXS (7.8±.17) were lower than either control males (9.2±.17) or females with FXS (8.8±.19).

**Figure 2.**
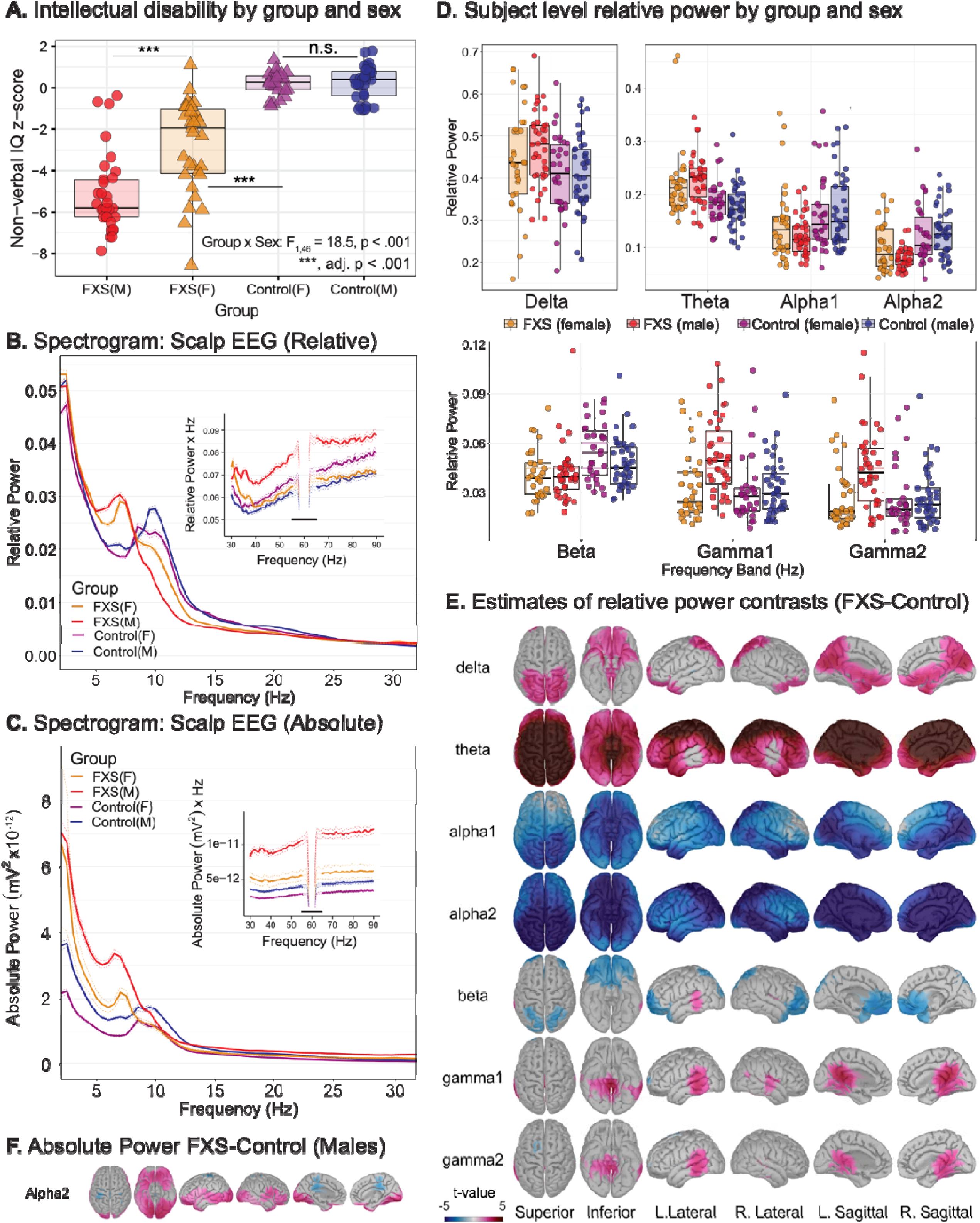
Spectral alterations are suggestive of neocortical hyperexcitability in FXS. Scalp and source-localization of dense array electroencephalography (EEG) suggest broadband power disruptions in frequency oscillations in FXS (n=70) compared to age- and sex-matched controls (n=71). **(A)** Subject-level scatter plot of intellectual disability as estimated by non-verbal intelligence z-scores (NVIQ) by allele group. NVIQ was markedly reduced in males with FXS and to a lesser degree in females with FXS. In neurodevelopmental research, NVIQ estimates general intelligence in populations that may not have verbal communication. **(B)** Mean relative scalp EEG power spectrogram (thick lines) with 95% confidence intervals (dotted tracings) by group and sex and inset of depicting gamma frequencies (30-90Hz). A marked leftward shift in dominant frequency in FXS groups and elevated gamma power. Is present. **(C)** Low-frequency changes in FXS groups persist in absolute power spectrograms and are not dependent on elevated gamma activity. **(D)** Boxplots displaying a median and interquartile range of relative power by frequency band (split into subplots by frequency band to optimize scale), averaged for each significant ROI from group-level t-maps. Subject level data is superimposed as a scatterplot. **(E)** Group-level t-maps depicting FXS – control, vertex-by-vertex relative power differences by frequency band superimposed on brain surface models. Warmer (FXS > Control) and cooler (FXS < Control) color scale represents significant t-values (non-significant values as gray). **(F)** Vertex-level comparison of source estimated absolute power in males with FXS confirms decreased alpha power is not dependent on gamma activity.

#### Scalp EEG Absolute Power

To ascertain if these changes in relative alpha and theta power were dependent on the proportion of gamma power, we next examined absolute power in which each frequency band is considered independently. *Scalp topography:* Group comparison of scalp topography demonstrated elevation of absolute power, except within the alpha range where FXS and controls were generally statistically similar, but there were patches of increased alpha1 and decreased alpha2 in FXS (**see Figure S1B**). *Scalp spectrogram and peak alpha frequency (PAF):* The absolute power spectrogram displayed an increase in FXS participants across most frequencies (**see Figure 2C**). However, a significant decrease in absolute alpha2 power is visible in males with FXS. Thus, participants with FXS (exemplified in males) show similar or lower levels of alpha2 power irrespective of activity in other frequency bands. The results of these analyses 1) confirm that relative power differences (especially within the alpha band) are not dependent on the proportion of gamma power and 2) the need for spectral normalization to account for large baseline differences in absolute power at the group level. Absolute power is more influenced by skull thickness and head geometry, which vary considerably across participants^20^, and such factors are present in FXS^21,22^. Thus, we primarily performed relative power normalization in subsequent sections to mitigate subject- and group-specific biases, evident in absolute power analyses.

### Use of source localization to resolve scalp-level findings

Though evidence from scalp EEG findings suggests cortical hyperexcitability, drawing conclusions regarding the spatial distribution of these changes is limited as electrode activity represents a mixture of underlying brain sources^23,24^. We employed source modeling for the first time in FXS to overcome scalp EEG analysis limitations and localize significant group differences by frequency band^25,26^. As predicted by the TCD model, we hypothesized that changes in low-frequency bands would be global, but increased gamma activity would be more localized. A depth-weighted minimum norm estimate (MNE) model based on the cortical envelope of the Montreal Neurological Institute (MNI) ICBM152 common brain template was used to perform source localization^27^. The result was a triangular mesh of 15,002 vertices, with each vertex representing a time series of current source density (CSD). Vertices were parcellated into 68 cortical nodes according to the Desikan-Killiany (DK) atlas^28^ for the remainder of the analyses. We also employed two top-level grouping strategies of nodes: 1) regional (region) in which adjacent nodes represent segments of cortical lobes and 2) resting-state networks (RSNs) in which nodes were arranged by previously characterized highly structured EEG functional dynamics^29^. A visual atlas of the nodes is presented in **Appendix 1**. Regions and RSNs are tabulated in **Appendix 2**. Thus, analyses were performed at either the vertex, node, region, or network level based on hypotheses.

### Source localization reveals a global decrease in alpha activity and localized changes in gamma power

#### Vertex Level

We first performed a high-resolution group comparison of the relative power of the cortical envelope at the vertex level. **Figure 2D and 2E** depict 5% FDR-corrected significance between-group contrasts (FXS-Controls). In participants with FXS, we observed a global reduction in alpha2 power and a global increase in theta power. In contrast, increases in gamma activity in FXS were primarily restricted to the bilateral temporal lobes and regions of the parietal and occipital lobes. We again confirmed that the decrease in alpha2 was not due to relative power normalization by comparing source-localized absolute power between groups. Even considering that mean absolute power was increased across all frequency bands in FXS, at the group level, alpha1 and alpha2 activity were higher only in lateral regions in FXS (**see Figure S2)**. In males with FXS, absolute alpha2 power had significant clusters of increased and decreased alpha2 absolute power (**see Figure 2F**).

#### Region Level

A recent study examining TCD signatures in Parkinson’s disease, neuropathic pain, tinnitus, and depression found spectrally equivalent but spatially distinct forms of TCD depending on the disorder^30^. After parcellation of nodes into cortical regions, we examined the group by sex differences of spectral activity by frequency band. Based on predicted lower or absent FMRP levels and higher burden of neuropsychiatric symptoms, we expected higher deviations of spectral power in males with FXS than controls. We found a robust interaction effect supporting our hypothesis between group, sex, frequency band, and region (F_78,66587_=1.64, p<.001). Results from pairwise comparisons of same-sex groups showed that participants with FXS had increased theta and decreased alpha power across cortical regions, but this was more pronounced among males with FXS (**see Table 2**). In addition, males with FXS, but not females with FXS, exhibited increased gamma power (**see Figure 3A**). Gamma power peaked across temporal regions but did not reach statistical significance in prefrontal (gamma1 and gamma2) or left occipital regions (gamma2).

**Figure 3.**
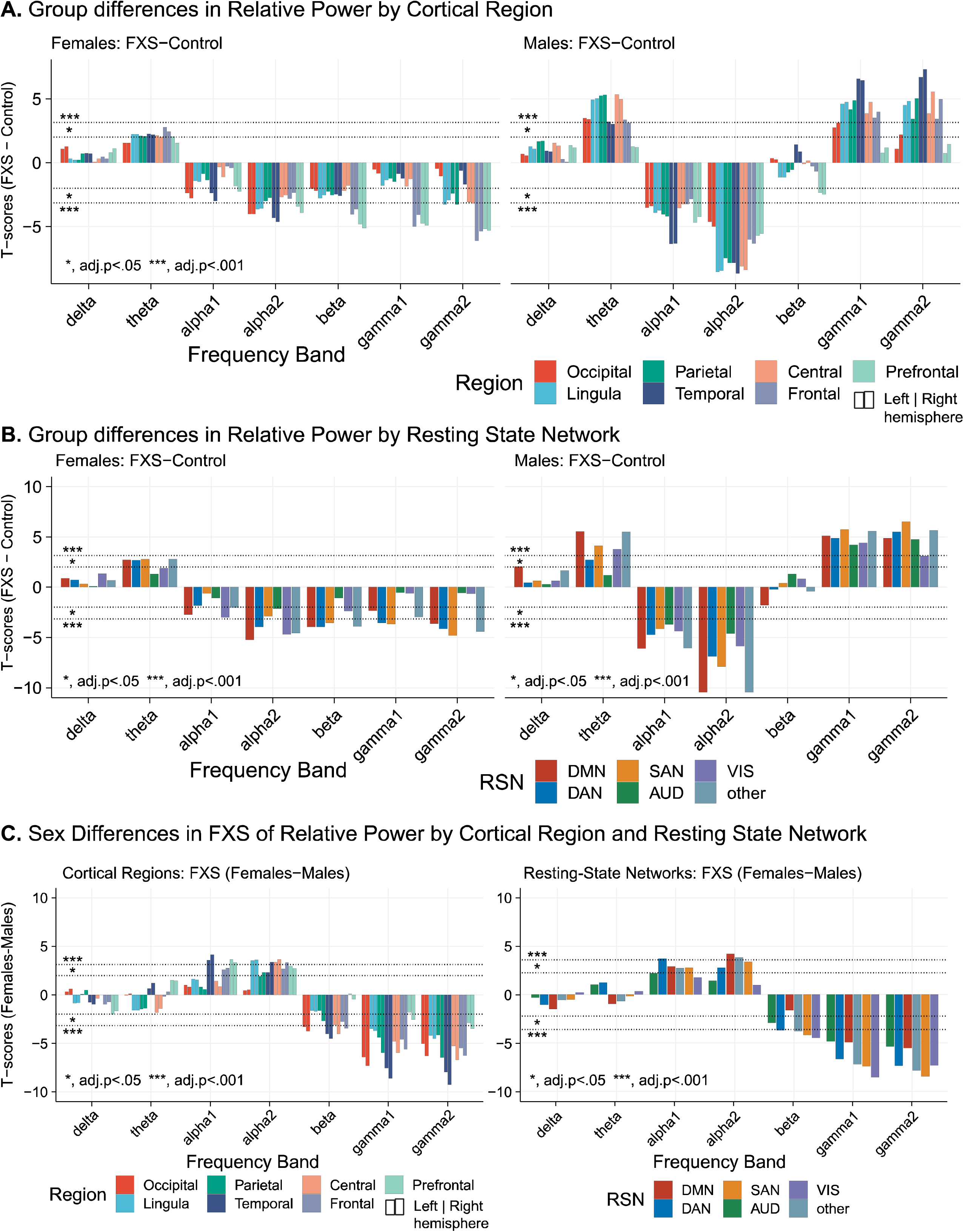
Relative power differences vary by group, sex, frequency band and by either cortical region or resting-state network (RSN). Bar plots represent 5% FDR corrected pairwise contrasts of model estimates of log relative power. For region plots, left and right bars with a color corresponding to right and left hemispheres, respectively. Positive t-values indicate that log relative power estimates in FXS are greater than those in control. (A) Sex-matched group differences in log relative power by cortical region. Males with FXS demonstrate region-specific increases in gamma power compared to controls with sparing of prefrontal regions. (B) Sex-matched group differences in log relative power across RSNs. A significant increase in theta and gamma power, as well as a decrease in alpha power, were observed across cognitive and sensory RSNs of males with FXS. Compared to control females, females with FXS had only modest changes in RSNs including similar gamma levels in visual and auditory networks. Obligate mosaicism in *Fmr1* in females with full mutation FXS may attenuate EEG alterations. We explored sex differences in relative power within the FXS group by region (C) and RSN (D). Positive t-values indicate that log relative power estimates in males with FXS are greater than females with FXS. There were fewer differences between males and females with FXS than in control comparisons, except that gamma activity remained elevated in most regions and all RSNs. Resting-state network abbreviations: DMN, default mode network; DAN, dorsal attention network; SAN, salient affective network; VIS, visual attention network; AUD, auditory network.

**Table 2.** Sex and Group Differences in Cortical Regions by Frequency Band. Estimated least-squared mean differences (FXS-Control) in log relative power per cortical region by frequency band denoted with 5% false discovery rate (FDR) statistical significance. FDR (5%) adjusted significant contrasts indicated with asterisks (*, p < .05; **, p < .001; ***, p < 1×10^−5^). See Appendix 2 for descriptions of nodes within each region.

| L/R | Region | Sex | Frequency Bands |  |  |  |  |  |  |
| --- | --- | --- | --- | --- | --- | --- | --- | --- | --- |
|  |  |  | delta | theta | alpha1 | alpha2 | beta | gamma1 | gamma2 |
| Left | Central | F | .003 | .20 | -.06 | -.18 | -.18 | -.14 | -.25* |
|  |  | M | .17 | .44*** | -.37*** | -.61*** | .02 | .39*** | .43*** |
|  | Frontal | F | .03 | .21* | -.05 | -.17 | -.24* | -.29*** | -.38*** |
|  |  | M | .06 | .29*** | -.30*** | -.48*** | -.04 | .28*** | .29*** |
|  | Lingula | F | .01 | .19 | -.11 | -.21* | -.20 | -.13 | -.26* |
|  |  | M | .12 | .38*** | -.36*** | -.58*** | -.06 | .36*** | .37*** |
|  | Occipital | F | .08 | .15 | -.18 | -.26* | -.16 | -.02 | -.01 |
|  |  | M | .14 | .28*** | -.36*** | -.42*** | -.03 | .21* | .10 |
|  | Parietal | F | .03 | .19 | -.10 | -.20 | -.19 | -.09 | -.18 |
|  |  | M | .19* | .39*** | -.39*** | -.54*** | -.04 | .34*** | .31*** |
|  | Prefrontal | F | .05 | .18 | -.12 | -.21* | -.31*** | -.33*** | -.39*** |
|  |  | M | .10 | .18 | -.34*** | -.46*** | -.12 | .15 | .14 |
|  | Temporal | F | .04 | .17* | -.13 | -.22** | -.15 | -.05 | -.06 |
|  |  | M | .09 | .28*** | -.39*** | -.49*** | .05 | .39*** | .38*** |
| Right | Central | F | .01 | .18 | -.09 | -.18 | -.16 | -.09 | -.26* |
|  |  | M | .15 | .42*** | -.36*** | -.62*** | .01 | .42*** | .50*** |
|  | Frontal | F | .005 | .19 | -.05 | -.15 | -.24* | -.25** | -.36*** |
|  |  | M | .04 | .29*** | -.29*** | -.50*** | -.02 | .33*** | .39*** |
|  | Lingula | F | .01 | .19 | -.11 | -.22* | -.19 | -.10 | -.24* |
|  |  | M | .11 | .38*** | -.36*** | -.58*** | -.06 | .37*** | .38*** |
|  | Occipital | F | .09 | .14 | -.19 | -.25* | -.16 | -.03 | -.04 |
|  |  | M | .14 | .28*** | -.36*** | -.43*** | -.03 | .23* | .17 |
|  | Parietal | F | .06 | .17 | -.13 | -.19 | -.19 | -.08 | -.21* |
|  |  | M | .19* | .39*** | -.40*** | -.56*** | -.05 | .37*** | .38*** |
|  | Prefrontal | F | .06 | .16 | -.14 | -.22* | -.32*** | -.32*** | -.38*** |
|  |  | M | .08 | .18 | -.31*** | -.45*** | -.09 | .20* | .20* |
|  | Temporal | F | .05 | .15 | -.15 | -.22** | -.16 | -.08 | -.12 |
|  |  | M | .09 | .26*** | -.38*** | -.52*** | .02 | .37*** | .41*** |

#### Network Level

Neuroimaging studies have identified modular brain networks related to higher-order cognitive, affective, and motor functions^31^. Thus, we extended our analysis to EEG-based RSNs, which represent functional networks rather than contiguous anatomical regions (**see Appendix 2**). The interaction of group, sex, frequency band, and RSN was a significant linear predictor of log relative power (F_30,66811_=2.21, p<.001; **see Figure 3B**). Females with FXS exhibited fewer and more modest changes from control females than their male counterparts. Females with FXS, for example, had similar gamma1 and gamma2 power across auditory and visual networks and decreased gamma power across cognitive networks to control females. In contrast, males with FXS demonstrated significant elevations in gamma power across all RSNs.

### Modulation of gamma power abnormalities by sex

We hypothesized that sex, a key determinant of phenotype in FXS^32,33^, is associated with stepwise variation in EEG changes. We examined the above models for within-group spectral power contrasts between males and females with FXS by region and network (**see Figure 3C**).

#### Region Level

No significant difference in theta power was found between males and females with FXS. Females with FXS generally displayed greater alpha1 and alpha2 power across regions, driven primarily by changes in the temporal, occipital, and frontal regions. Gamma1 and gamma2 power were highly elevated in males with FXS across most cortical regions (adj. p<.001) with the greatest difference in the temporal regions and modest differences in the prefrontal regions. <u>Network Level:</u> No significant differences in theta power were found between males and females with FXS. Interestingly, no sex differences were found from alpha1 and alpha2 power across the two sensory RSNs (visual and auditory). Gamma1 and gamma2 power, however, was significantly increased in males with FXS across all RSNs (p<.001).

Overall, males with FXS appeared to have modest or no change in low-frequency power profiles than females with FXS, however, displayed pervasive elevation of gamma power.

### Peak alpha frequency at source level reveals reduced frequency and loss of posterior-anterior configuration

#### Region Level

Reports of TCD have observed alterations in alpha activity including 1) leftward shift of PAF towards the theta frequency and 2) shifted spatial distribution (known as “anteriorization”) of “slow” alpha frequencies. Our goal was to determine how group, sex, and posterior-to-anterior cortical regions (occipital, parietal, central, and frontal) effect source localized PAF. A three-way interaction was present (F_3,4354_=4.303; p=0.005) suggesting a linear relationship between PAF and the interaction of group, sex, and region. *Frequency reduction of source PAF in FXS:* To confirm the presence of reduced or “slowed” PAF in FXS, we examined univariate pairwise contrasts between PAF within each posterior-anterior region between sex-matched groups. Both males and females with FXS had reduced PAF compared to controls, with the largest decrease in the parietal and central regions of males with FXS (**see Figure 4A**). *Spatial configuration of source PAF is lost in FXS:* We hypothesized that PAF would be reduced and lose its characteristic asymmetrical posterior-anterior topography, as with other TCD syndromes. In male and female controls and to a lesser extent females with FXS, PAF was peaked over parietal and central regions (**see Figure 4B**). In contrast, males with FXS displayed a relatively flat profile of PAF across posterior to anterior regions. PAF in males with FXS was mildly depressed from occipital to parietal regions and not significantly different between posterior and central regions.

**Figure 4.**
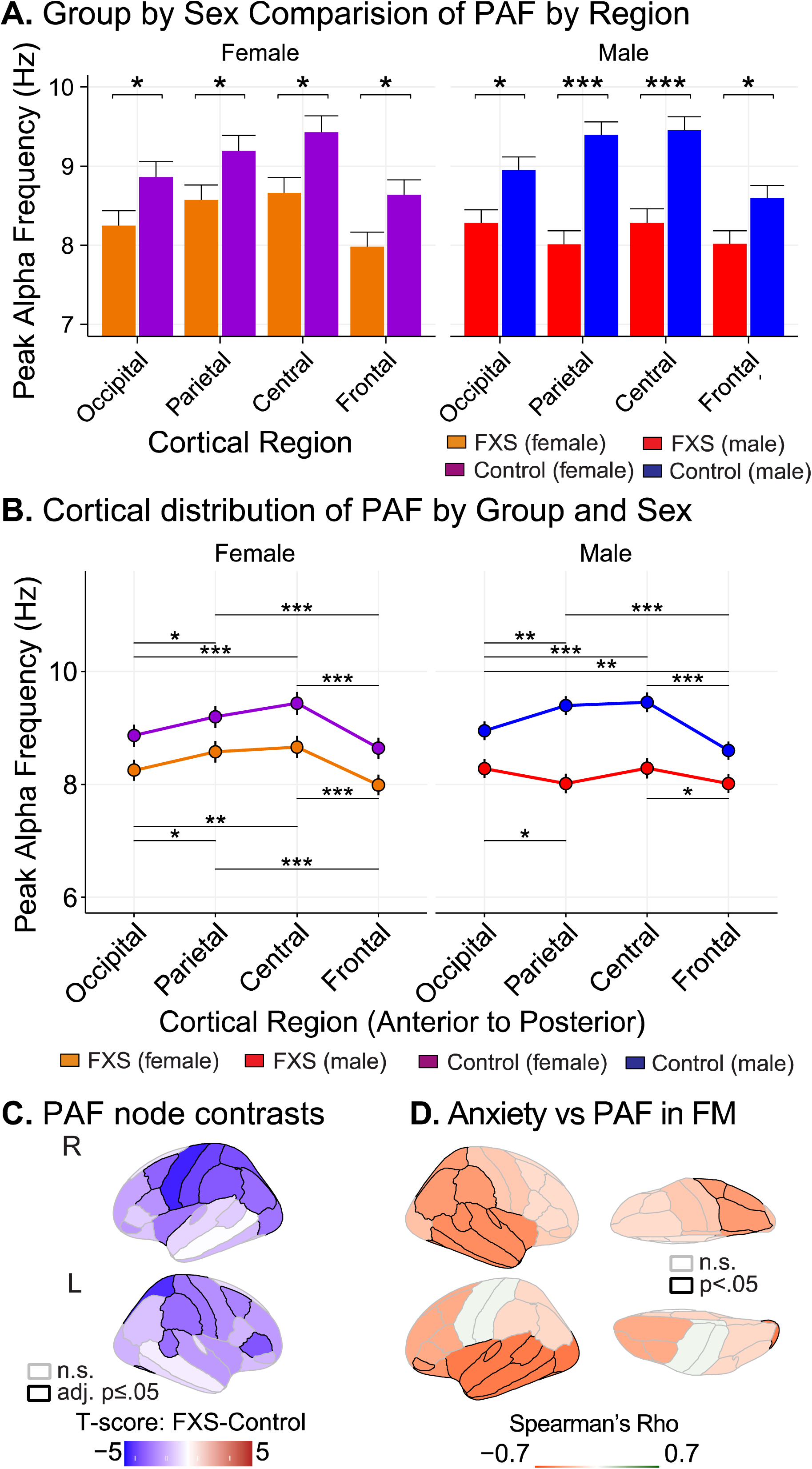
Alterations in peak alpha frequency (PAF) are supportive of thalamocortical dysrhythmia in FXS patients. The slowing of alpha oscillations has been observed across scalp potentials and intraoperative recordings and is suggestive of abnormalities in thalamocortical activity in several neuropsychiatric conditions. **(A)** Bar plots (mean ± standard error of least-squared mean estimates) comparing regional PAF by groups matched by sex. Participants with FXS have slower PAF across all cortical regions from controls. **(B)** Line plots comparing the within-subject distribution of PAF across posterior-anterior cortical regions. Males with FXS have a relatively even distribution of PAF across the posterior-anterior axis with no prominent central elevation as seen in matched controls. **(C)** Visualization of mean FXS-control differences of PAF by cortical node. Participants with FXS have broadly reduced PAF with the most prominent reductions in central and parietal nodes. See Appendix 1 for node atlas. **(D)** The severity of anxiety is inversely associated with PAF. Visualization of cortex plotting age-corrected Spearman correlations in full-mutation, non-mosaic males (FM; n=27; r(25)=-.41 to - .53, p ≤ .05). FM, full mutation FXS, non-mosaic males; LO, left occipital. Horizontal black bars: FDR-adjusted, post-hoc testing; *, adj. p ≤ .05; **, adj. p ≤ .01; ***, adj. p. ≤ 1×10-5.

#### Node Level

Given differences along the posterior-anterior axis, we next examined contrasts in PAF in all 68 atlas nodes to further clarify the spatial distribution of PAF reduction (**see Appendix 2**). Interestingly, we found a significant interaction effect between group and node (group x node; F_67,9378_=1.44, p=0.01), but no effect of sex. In approximately 51% of nodes (35/68), participants with FXS had a significantly lower PAF than controls (**see Table S3**). An atlas-based visualization is presented in **Figure 4C** which depicts t-values of the FXS-control contrast. The greatest difference in PAF between FXS and controls were found in the left parietal region including the supramarginal gyrus (F_140_=-4.7, 5% FDR <.001), inferior parietal gyrus (F_140_=-4.4, 5% FDR <.001) and precuneus (F_140_=-4, 5% FDR <.001).

### Theta, not alpha, power is predominantly associated with gamma power in FXS

Power-power CFC is the association (in normalized Fisher’s Z-transformed correlation coefficients) between time series of EEG power between two frequencies ^34^. In TCD syndromes, theta power is more strongly correlated with gamma power than is alpha power. These alterations have been associated with disruptions in the task-related functional activity and cognitive processing^35^. We evaluated the effects of group and sex on theta-gamma1, alpha1-gamma1, and alpha2-gamma1 CFC. As alterations of low-frequency power in FXS occur globally, we specifically examined whole-brain theta, alpha1, and alpha2 CFC with node-level and network-level gamma1 activity.

#### Node CFC

We first examined the effect of the lower frequency band (theta, alpha1, or alpha2), diagnostic group, sex, and cortical node on gamma1 CFC. Unexpectedly, sex did not have a significant main or interaction effect and was not retained in the final model. We found that lower frequency band, group, and cortical node had a significant interaction effect on gamma1 CFC (F_134,28217_=1.93, p<.0001). Participants with FXS demonstrated an inversion of CFC relationships such that increases in theta power, not alpha, were associated with decreased gamma1 power (**see Figure 5A and 5B**). This increase was particularly manifest over cingulate, temporal, and parietal nodes (**see Table S4**). In contrast to controls, increased alpha2 power was directly associated with increased gamma power.

**Figure 5:**
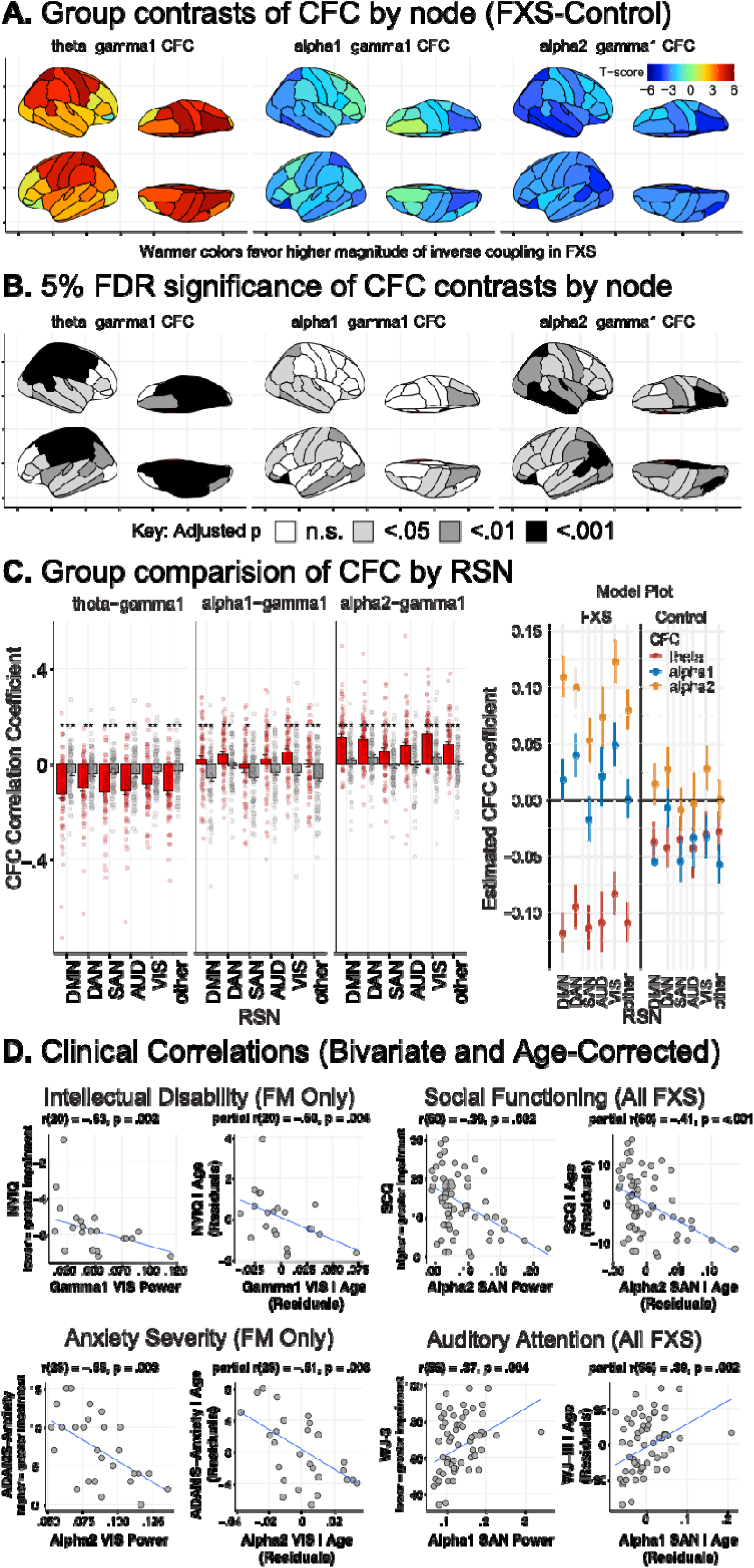
The power-power cross-frequency coupling (CFC) between theta power and gamma power in TCD syndromes is stronger than that between alpha power and gamma power. (**A**) Brain atlas heatmaps depict group-level differences (t-values) between CFC. Since CFC coefficients can represent either a direct or inverse relationship between frequencies, higher the t values, the greater the difference in the magnitude of the coupling. Participants with FXS showed a significantly higher magnitude of inverse theta-gamma1 CFC compared to controls, especially over the parietal and central regions. (**B**) Brain atlas plots with shaded areas representing 5% FDR-corrected significance of pairwise contrasts. (**C**) Left: Barplots of mean ± standard error of least-squared mean estimates (with superimposed subject-level data) of CFC by RSN (Left). Right: Mean model plot of group by CFC type across RSN demonstrating a prominent increase in inverse theta-gamma1 CFC and increase direct alpha2-gamma CFC in the FXS group. See Appendix 1 for node atlas. (**D**) Exemplar clinical correlations with EEG variables. Scatterplots in each quadrant depict subject-level bivariate (left) and partial (right; age-corrected) Spearman’s correlations. Left: Intellectual function and anxiety are significantly correlated with gamma1 and alpha2 power, respectively, in full mutation, non-mosaic males (FM). Right: Across all FXS subjects, social functioning and auditory attention were significantly correlated with alpha power. Resting-state network abbreviations: DMN, default mode network; DAN, dorsal attention network; SAN, salient affective network; VIS, visual attention network; AUD, auditory network. FDR-adjusted p values, *, adj. p < .05; ***, adj. p. < 1×10^−5^.

#### Network CFC

To better understand the functional impact of altered node-level CFC, we also compared CFC measured across RSNs. We found a similar trend to our individual node analysis, with a significant interaction effect of the lower frequency band, group, and RSN (F_10,28589_=3.02, p<.001). Across all functional networks, contrasts of model estimates revealed that participants with FXS had a greater magnitude of inverse theta CFC and increased alpha2 CFC with gamma1 (**see Figure 5C and Table S5**).

### Clinical Correlations

Due to the broad age range of the sample, partial Spearman’s correlations were conducted to control for effects of age on findings between EEG features and clinical measures. We first examined 5% FDR corrected correlations between (1) spectral power (**see Table S6-S9**), (2) PAF (**see Table S10**), and (3) CFC (**see Table S11**) values across region and RSN for all participants with FXS. FDR correction was performed on the entire correlation table for each of the three main EEG features: source estimated band power, PAF, and CFC. We next conducted uncorrected exploratory correlation analysis on a subgroup of full-mutation, non-mosaic males (FM) to eliminate confounding with potential mosaic effects and examine findings in a more homogenous group that parallels *Fmr1*^-/-^ KO mouse. We have hosted an interactive statistical R web application (https://epedapati.shinyapps.io/tcdfxs_corr/) for review and visualization. See Methods for abbreviation key for cortical regions and RSNs.

#### Intellectual Quotient (IQ)

*All FXS:* After FDR adjustment, there was a trend-level association such that increased verbal IQ (VIQ) was associated with elevated PAF (VIQ; SAN: r(62)=.39; p=.002, adj. p=.081). *FM subgroup:* Reduced NVIQ was inversely associated with elevations in gamma power across multiple regions and RSNs (NVIQ; DMN, DAN, AUD, VIS, Temporal, Parietal, Occipital: r(20) = -.44 to -.60, p≤.05; **for exemplar see Figure 5D top left**). CFC demonstrated significant correlations with NVIQ and VIQ. A scatter plot is helpful in interpreting CFC to represent the direction of the relationship. A larger relationship between increased theta and gamma1 power (theta-gamma1 CFC) was associated with increased NVIQ (SAN, DAN: r(20)=.44 to .49, p≤.05). Unexpectedly, increased association of alpha1 (alpha1-gamma CFC: DAN, AUD, VIS: r(20)=.44 to .53, p≤.05) or alpha2 (alpha2-gamma CFC: DAN: r(20)=.46, p≤.05) with gamma1 power was associated with increased NVIQ as well. For VIQ, greater inverse alpha-gamma CFC (e.g., higher alpha power correlated with decreased gamma power) was correlated with higher verbal ability (alpha1-gamma1: LT: r(20)=-.56, p≤.05; alpha2-gamma1: LT: r(20)=-.56, p≤.01).

#### Anxiety and OCD

The Anxiety, Depression and Mood Scale (ADAMS) anxiety and obsessive subscales assess the burden of these symptoms in individuals with intellectual disability^36^. *All FXS:* Higher levels of obsessive-compulsive symptoms were associated with lower alpha1 and alpha2 power across most cortical regions and RSNs (ADAMS-OCD: r(62)=-.38 to -.52; adj. p≤.05). *FM subgroup:* Higher anxiety levels were associated with lower alpha1 (RPF, RT; r(25)=-.45 to -.48; adj. p≤.05) and alpha2 (RPF,LPF,RT,LT,RO,LO, and VIS; ADAMS-Anxiety r(25)=-.48 to -.60; p≤.01 to .05; **for exemplar see Figure 5D bottom left**) levels across several regions and within the visual network. Higher anxiety levels were associated with lower PAF across multiple regions and the DMN (RPF, LPF, RT, LT, LO, RP, and DMN: r(25)=-.41 to -.53; p≤.05). Higher anxiety scores were associated with increased alpha1-gamma1 CFC (RO, VIS; ADAMS-Anxiety: r(25)=.41-.44; p≤.05). Similar directional effect was present for obsessive-compulsive symptoms with theta-gamma1, alpha1-gamma1, and alpha2-gamma CFC (RO, VIS; ADAMS-OCD: r(25)=.39-.40; p≤.05).

#### Behavioral Symptoms

Aberrant Behavior Checklist (ABC) is a widely used measure in ASD and FXS studies where higher score indicate more severe symptoms across behavioral domains ^37^. *All FXS:* Alpha power was inversely correlated with the abnormal speech subscale (prefrontal, SAN; r(59)=-.38 to -.48, adj. p≤.01 to .05), hyperactivity (AUD, DAN, PF, LT, RT; r(59)=-.40 to - .49, adj. p≤.05), and stereotyped behaviors (PF, RT, LT, RO, LO, DMN, DAN, and VIS; r(59)=-.39 to -.50, adj. p≤.05). *FM subgroup:* Elevations of gamma power were significantly associated with increased severity in behavioral domains including abnormal speech (RPF, LPF, RT, RP, RL, LL, DMN, DAN, SAN, AUD; r(22)=.36 to .57; p≤.05), hyperactivity (LF, RP; r(22)=.42 to .43, p≤.05), irritability (RP r(22)=.42, p≤.05), and lethargy/social withdrawal (LF; r(22)=.43, p≤.05). Reduced alpha activity was associated with worsening symptoms including abnormal speech (alpha1: LPF, RPF; r(22)=-42 to -.49, p≤.05), stereotypy (alpha1: LPF, RO, RPF; r(22)=-.43 to - 66, alpha2: LPF, RPF; r(22)=-.48 to -.55), lethargy (alpha1: LPF, RPF; r(22)=-.45 to -.51, p≤.05, alpha2: LPF, RPF; r(22)=-.44 to -.49, p≤05).

#### Social Communication

The SCQ is a brief screening instrument to evaluate social-communication skills across the lifetime where higher values indicate greater impairment ^38^. *All FXS*: Lower levels of social-communication functioning (higher SCQ score) were associated with decreased alpha1 (LPF, RPF, LT, RT, DMN, DAN, SAN, and VIS; r(60)=-.39 to -.41; adj. p ≤ .05) and alpha2 power (RPF, LPF, RC, LF, RL, LL, DMN, DAN, and SAN; r(60)=-.37 to -.52; **for exemplar see Figure 5D top right**). *FM subgroup*: Lower levels of social-communication functioning were associated with alpha1 (RPF; r(22)=-.43; p≤.05) and alpha2 (RPF; r(22)=-.46; p≤.05).

#### Auditory Attention

Selective auditory attention (including speech-sound discrimination and resistance to auditory-stimulus) was measured using the Woodcock-Johnson III auditory attention task (WJ3)^39^. *All FXS*: We found that improved auditory attention performance was associated with alpha1 power (SAN; r(56)=.39, adj. p≤.05; **for exemplar see Figure 5D bottom left**). *FM subgroup*: Reduced auditory attention performance was associated with decreased alpha1 (LO; r(21)=.52; p≤.05), but increased theta (LO, RO, LP, and RP; r(21)=.45 to .52, p ≤ .05), gamma1 (LO, RP, and VIS; r(21)=-.42,p≤.05), and gamma2 power (LO, RO, RP, and VIS; r(21)=-.44,p≤.05).

## Discussion

In the present study, we incorporate several novel findings derived from resting-state EEG to provide a unifying thalamocortical model to explain alterations of neural activity in FXS. The findings provide spatial and functional context to previously reported scalp-level EEG abnormalities in alpha/theta oscillations and elevated non-myogenic gamma activity. First, we observed clinically associated increases in gamma activity which varies by sex. Second, we report global changes in alpha and theta power, including a reduction and altered spatial configuration of PAF. Third, we demonstrate a predominance of regional and RSN-based theta-gamma CFC in FXS. These results were obtained from a well-powered sample of individuals with FXS and age- and sex-matched controls, using source modeling to identify effects with anatomical and functional ROIs. Our results highlight system-level features to enhance the development of patient-oriented biomarkers and provide key physiological insight into the neural activity of a prototypical monogenetic NDD^40^.

### Evidence of TCD in FXS

The present results suggest that altered thalamocortical modulation may be a key mechanism of neocortical hyperexcitability in FXS and encourage further investigation of thalamocortical physiology in FXS. In FXS, thalamic abnormalities have been previously reported in neuroimaging studies, including lower fractional anisotropy between thalamus and neocortex^41^, reduced thalamic grey matter density^42^, reduced thalamic GABA-A receptor density^43^. However, to date, the functional and physiological impact of these changes were not well understood. Thalamocortical structures are interconnected by widespread excitatory connections, in which abnormalities have been associated with cortical hyperexcitability, including epilepsy^44^. The role of the thalamus has also continued to evolve from a “relay station” into a dynamic center for contextual modulation of cortical circuits^45^. Though large-scale direct measurements of thalamic contributions to neocortical excitability in FXS are unlikely, previous invasive experiments in humans, such as with stereotaxic EEG, have bolstered confidence that surface EEG oscillations are a proxy for thalamocortical activity^46^. In this sense, the observed EEG alterations including alterations associated with TCD may reflect the functional significance of the previously reported thalamic findings in FXS.

### Localized increases in gamma power

EEG is an ideal method for studying real-time brain activity at high frequencies, but the addition of source modeling can dramatically improve spatial resolution at the cortical surface^26^. Increases in gamma power were localized primarily to the temporal cortices and portions of the parietal and occipital lobes. Gamma oscillations hold a special interest in NDDs because of their relation to cortical excitability^5,47^, association with cognitive processes^48^, and analogous measurability in animal models^6^. The role of gamma oscillations is increasingly nuanced, such that precise synchrony in gamma activity is contributory to higher-order cognition^49,50^, but also that a modest degree of asynchrony or “noise” represents physiological processes^51-53^. Nevertheless, asynchronous (usually broadband) gamma power, above what is typically expected, has been associated with disease states^48^ as well as with reduced spike precision and spectral leakage of spiking activities in microcircuit preparations^54^.

### Variability in gamma power is associated with sex

Gamma activity varied significantly based on sex and was highly predictive of core cognitive and neuropsychiatric symptoms in FXS. As sex is highly predictive of phenotypic differences in FXS^32^, we also expected sex to have a large effect on EEG activity. Unexpectedly, we found females (who are obligate mosaics) generally demonstrated similar low-frequency alterations from controls as males with FXS, however, did not share similar profiles of gamma power. Indeed, when compared to female controls or males with FXS, females with FXS on average showed either no difference or a modest *reduction* in gamma power. Based on these findings, FMRP expression is associated with variability in gamma power, however the underlying local and system-level mechanisms leading to these changes are unknown. Within-group sex differences suggest that full-mutation females, who likely express more FMRP than their male counterparts, may moderate neocortical hyperexcitability as assessed by background gamma activity. Indeed, we found increased gamma activity was a predictor of the severity of abnormal speech in both sexes, and among the full-mutation, non-mosaic male subgroup increased gamma activity was predictive of worsening of abnormal speech, hyperactivity, intellectual function, and auditory attention features.

### Marked reduction in alpha activity, including PAF, in FXS

The global increase in theta power, decrease in alpha power, and marked reduction of PAF into the theta band implicate changes in thalamocortical circuits in FXS. Previously it was assumed that alpha generators resided in the thalamus, but recent research from human invasive recordings have placed new emphasis on cortical generators, while still confirming the central role of the thalamocortical system in driving alpha activity and PAF^46,55^. Specifically, a PAF between 10-13 Hz appears to have special relevance to functional brain physiology^49,56-59^. Like a radio receiver that can only tune specific electromagnetic frequencies, neurons and oscillatory networks appear to also demonstrate a frequency preference^60,61^. The “switch” to a dominant peak rhythm at 4-8 Hz in FXS rather than operating peaks closer to 10-13 Hz, may be insufficient to drive neural ensembles with an alpha preference^57,59,62,63^. Conversely, as theta dominance increases, alpha activity may lose its canonical role as an inhibitor of the circuits that provide optimal time for processing sensory and cognitive information^64,65^.

### Clinical relevance of changes in low-frequency oscillations

Theta and alpha oscillations have been implicated in different aspects of cognitive control, for example^66^. Not surprisingly, across the FXS cohort, diminished alpha power and PAF was associated with a broad spectrum of neuropsychiatric features common in FXS, including auditory attention, hyperactivity, stereotyped behavior, obsessive-compulsive behavior, and social communication skills. Interestingly, alpha power (or PAF) was not significantly associated with cognitive scores across FXS, despite higher PAF corelating with higher intelligence in control populations^67^. Hence, it appears that subsets of neuropsychiatric symptoms may be associated with global changes in alpha activity, whereas other disorder features such as cognition may be associated with gamma disturbances. We speculate, since the normalization of gamma activity is associated with increased FMRP expression, the pattern of EEG correlations may reflect distinct mechanisms which underlie the variety of cognitive and behavioral phenotypes found in FXS. It may be possible to further explore these hypotheses in future studies which correlate specific EEG findings with domain-specific behavioral tasks.

### Predominance of theta power-power CFC in FXS

As in other examples of TCD, we found that theta-gamma CFC, rather than alpha-gamma CFC, is predominant in FXS compared to controls^68^. We did not find any effect of sex in our final model, which likely reflects the similar low-frequency alterations in males and females with FXS, despite relative differences in gamma activity. It has been proposed that in TCD, theta modulation does not produce the same lateral inhibition of gamma activity as alpha oscillations. This leads to a net increase in asynchronous, or background, gamma activity and spurious neural activity ^16^. Exploratory correlations in full mutation, non-mosaic males revealed significant associations between cognitive and neuropsychiatric feathers with CFC, but the interpretation of these findings remains complex and will require future experiments to parse. For example, a stronger inverse theta-gamma CFC relationship was associated with reduced cognitive scores, but we cannot infer either directionally or directly compare with other frequency-specific CFC in the same subject. However, the results provide a rich starting point for developing hypotheses to for future connectivity and causality analyses.

### Theoretical considerations of TCD in FXS

As the TCD model has not previously been explored in FXS, considering theoretical framework from other disorders associated with TCD can provide future direction. In tinnitus, where abnormal sensory perception has been consistently linked with slow-wave oscillations, it has been hypothesized that decreased organized input to the thalamus (i.e., deafferentation or tonic inhibition) leads to excessive theta band activity. When this shift occurs, inappropriate activation of the sensory cortices (reflected by increased gamma activity) results in the perception of tinnitus. Though not directly measured in humans, neocortical changes in the *Fmr1*^*-/-*^ KO mouse include intrinsic hyperexcitability of pyramidal neurons^3^ and fast-spiking parvalbumin (PV) cell dysfunction^69^ which is critical to “sharpening” synchronous neuronal activity^70,71^. Could changes leading to a “noisy” cortex disrupt organized feedback to the thalamus, and thus, thalamocortical signaling, result in alterations observed with EEG? The extent to which noninvasive approaches such as pharmaceutical challenge and perturbation studies can answer these questions remains an active area of study. It is not possible from the current data to determine if the measured changes, which suggest neocortical hyperexcitability, are compensatory or causative, so back-translational approaches are necessary to uncover the underlying mechanisms of these biomarkers. For example, in some *Fmr1*^-/-^ KO circuitry, compensatory mechanisms may partially restore global homeostasis^47^. Placed in a larger context, the circuit changes observed in FXS appear to disrupt more specialized circuits for higher-order cognition, emotional regulation, and sensory processing^72^, rather than to a level representing a common cause of epilepsy (which is rare and mild in FXS). Thus, subpopulations in FXS with residual *Fmr1* expression may vary in phenotype based on the extent of protein deficiency, protein distribution, developmental period, circuit function, and neuronal type.

### Limitations

Rather than assess detailed point-to-point connectivity, we used CFC to assess whole-brain averaged alpha or theta power to gamma power across individual cortical nodes to compare to reports of TCD in other disorders^12^. Within the scope of TCD, phase-amplitude (or phase-phase) coupling is not well-understood, but CFC is well-suited to answer hypotheses derived from our main analyses and compare to reports of TCD in other disorders. Despite ascertaining mosaic status in males, males with mosaicism are a relatively small subset (n=12) and underpowered for subgroup analysis. Additionally, males with mosaicism can vary phenotypically based on mosaic type (repeat number or methylation), further emphasizing the importance of a well-powered sample^33^. Although the effect of non-epileptic medications on the results cannot be ruled out, a medication naïve sample would preclude the inclusion of more severely affected individuals given the high rate of medication use in FXS ^73^. Previous EEG studies of medication effects in psychiatric populations^74^, including our own observations in FXS^75^, do not suggest effects as we have observed.

### Conclusion

In summary, we have identified several novel alterations in brain activity in a large sample of participants with FXS which display strong clinical associations. Our data from females with FXS is highly novel and provides evidence for reduced broadband gamma alterations but similar alterations in thalamocortical circuitry as in males. Ultimately, the changes we have observed may contribute to and maintain abnormal cortical states that reduce functional brain connectivity and regional function necessary for optimal brain functions. The findings are important in establishing a robust translational strategy for developing and testing new treatments with electrophysiological biomarkers that can transfer directly from the mouse model to patient studies.

## Methods

### Participants

The dataset included a total of 145 participants drawn from a large federally funded human neurophysiology study in FXS (National Institutes of Mental Health U54 HD082008). Exclusion criteria for FXS (confirmed by Southern Blot and polymerase chain reaction) participants included present history of unstable seizures (any treated seizure within one year) and scheduled use of benzodiazepines. Controls did not have treatment for neuropsychiatric illness as reported via clinical interview. All participants provided written informed consent (or assent as appropriate) prior to participation as approved by the institutional review board of Cincinnati Children’s Hospital Medical Center. Following blinded preprocessing, three recordings were discarded from further analysis due to excessive line-noise artifact (1 FXS, 2 controls) and one due to insufficient data due to intolerance of the EEG procedure (1 FXS). The final dataset consisted of 70 participants with a genetic diagnosis of full mutation FXS (Mean age= 20.5, SD=10; age range: 5.9-45.7; 32 females; 12 males with mosaicism) and 71 controls (Mean age= 22.2, SD= 10.7; age range: 5.9-48.2; 30 females). Females with full mutation FXS were included in the primary analyses and effects were confirmed in supplemental analyses of male participants. Age effects were examined in each model for significant fixed effects. Thirty-five FXS patients were on antidepressants and 18 were receiving atypical antipsychotics. These and other concurrent medications were only permitted if participant was on stable dosing for at least 6 weeks.

### Data Acquisition and Preprocessing

Five minutes of continuous EEG data was collected while participants were seated comfortably while watching a silent video (standardized across participants) to facilitate cooperation as in previous studies^12^. Recordings were collected at 1000 Hz sampling rate with an EGI NetAmp 400 with a 128-channel HydroCel electrode net (Magstim/EGI, Eugene, OR). Preprocessing: All data was blinded and coded regarding group, participant, or collection date. Data was exported in EGI raw format and imported into EEGLAB SET format in MATLAB (version 2018b, The MathWorks Inc., Natick, MA, USA). Raw EEG data was filtered using EEGLAB 14.1.2^76^ with a 2 Hz high pass digital zero-phase filter and a 55 to 65-Hz notch filter (with harmonics removed up to Nyquist frequency of the original sampling rate) to remove line noise. Raw data was visually inspected by an assistant who excluded segments of data with large amount of movement artifact and interpolated bad channels (no more than 5% per subject) using spherical spline interpolation implemented in EEGLAB 14. Data was average referenced. An artifact subspace reconstruction (ASR) approach was carried out with the “clean_rawdata” function (with default parameters) to repair data segments of artifact by applying a reconstruction mixing matrix from non-interpolated neighboring channels. The mixing matrix is computed from clean segments from within the EEG data^77^. Blind source separation was performed with temporal Independent Component Analysis (ICA) on each preprocessed dataset using the extended INFOMAX algorithm^78,79^ with PCA rank reduction (further reduced for interpolated channels). This approach was recently validated to effectively reduce myogenic contamination from approximately 25-98 Hz^80^. Resulting components were manually reviewed and categorized for eye movement/blinks, muscle movement, channel noise, or cardiac artifact based on temporospatial and spectral features and back projected to remove artifact. Resulting non-artifactual independent components are near-independent in time course activity and resemble dipolar scalp projections and have been proposed to represent spatially coherent local field activity within a single cortical area^81^. Data was divided into 2-second epochs and manually reviewed for noise artifacts. Summary of artifact cleaning is presented in Supplemental Table 2 and demonstrate no significant differences between preprocessing measures between groups. Raw EEG data is available to the public as federally mandated at the National Database for Autism Research (NDAR).

### Source Modeling

Minimum norm estimation (MNE) is a widely adopted solution to the inverse problem in which current estimates are calculated at every spatial location in source space to minimize the total power across the cortex^82^. Thus, MNE models, in contrast to dipole fitting, produce uniform maps across subjects which is well-suited for group comparisons and can provide resolution comparable to magnetoencephalography (MEG)^83^. For each subject, the first 80-s of artifact-free data from each of the EGI 128-channel electrodes were co-registered with an Montreal Neurological Institute (MNI) averaged ICBM152 common brain template^84^. The degree of accuracy and precision of EEG source localization is debated, but intracortical recordings during epileptic surgery^85^, surface and deep brain stimulation^86^, and comparisons with functional magnetic resonance imaging (fMRI)^87^ estimate focal localization at 1.5 cm for superficial neocortex. Thus, even with standard head models and spatial smoothing EEG is suitable for studying high-frequency brain activity in vivo clinical studies^88^. Open M/EEG^89^ was used to compute a 15,000 vertices lead-field mesh incorporating electrode distances. Noise covariance was set as an identity matrix as recommended for scalp resting EEG recordings^27^. Construction of L2-normed, depth-weighted MNE source model to generate a current source density (CSD) map (units: picoampere-meter) was performed in Brainstorm^27^ and used to reconstruct time series activations at each vertex.

### Anatomical and Functional Parcellation

Individual vertices from the lead-field mesh were grouped into 68 cortical nodes according to the Desikan-Killiany (DK) atlas^28^. We opted to study both an anatomical and functional configuration of the atlas nodes. The anatomical configuration was derived from the DK atlas and consisted of categorizing the 68 notes into 14 regions: occipital (O), lingual (L), parietal (P), temporal (T), central (C), frontal (F), and prefrontal (PF) each with a right (R) or left (L) designation. Functional source EEG resting-state networks have been derived from examining their dynamic properties and similarities to networks identified by other neuroimaging techniques (diffusion tensor imaging, fMRI, and magnetoencephalography)^29^. Following this template, we assigned 44 cortical nodes into resting state networks including default mode network (DMN), dorsal attention network (DAN), salient affective network (SAN), auditory network (AUD), and visual network (VIS). Remaining nodes not associated with a functional network are classified by convention as “other”.

### Spectral Power

For all analyses, we divided spectral power into 7 frequency bands: delta (2–3.5Hz), theta (3.5– 7.5 Hz), alpha1 (8–10 Hz), alpha2 (10–12.5 Hz), beta (13–30 Hz), and gamma1 (30–55 Hz), and gamma2 (65-90 Hz). Upper alpha bands are associated with more complex cognitive processing^65,90^, and lower alpha bands have been primarily associated with attentional processes including alertness, expectancy, and vigilance^91^.

### Scalp EEG

Segmented data (2-s) from 108 scalp EEG channels were detrended, tapered with a Hanning window, and transformed into Fourier coefficients representing 0.5 Hz frequency steps. Fourier coefficients were squared to compute absolute power and divided into frequency bins. Relative power was defined as the band-specific cumulative absolute power (V^2^/Hz) divided by the total power across all defined bands and then averaged over available trials. For source data, Welch’s method (50% overlap with Hanning window) was used to estimate spectral power from each vertex CSD time series. To facilitate group comparisons, we used a circularly symmetric Gaussian smoothing kernel with a full width half maximum (FWHM) size of 3 mm^92^ across all vertices. Relative power calculations were performed identically to scalp electrode power.

### Peak alpha frequency

The average dominant frequency (i.e., alpha peak) was determined by the “findpeak” function in MATLAB to identify the frequency of the maximum absolute logarithmic power between 6-14 Hz from each channel or DK node spectrogram^93^.

### Clinical Measures

Stanford-Binet Intelligence Scale 5th Ed. (SBS)^94^ was conducted by trained clinicians in both FXS and control participants. Due to floor effects, deviation IQ scores^95^ were computed to capture variability in cognitive functioning. Assessments were completed by the primary caregivers for FXS patients including the Social Communication Questionnaire (SCQ)^96^, Anxiety, Depression, and Mood Scale (ADAMS)^36^, Woodcock-Johnson III Tests of Cognitive Abilities, Auditory Attention subscale (WJ3)^39^.

### Statistical analysis

Statistical analysis was performed with MATLAB 2018b (MathWorks, Natick, MA, USA), SAS 9.4 (SAS Institute Inc., Cary, NC, USA), and R (4.1, Vienna, Austria).

### Power and Sample Size

Differences in gamma1 power in FXS compared to controls in previous studies have effect sizes from .63 to 1.75, similar to effect sizes in prior studies of N1 amplitudes in FXS^12,13,75,97^. Based on these effect sizes, comparing 70 FXS patients (50% males) and 70 TD controls provides power to detect the primary EEG outcome with approximately power > .90 (using an omnibus F-test with an alpha of .05). In line with reproducible research guidelines, scripts for generation of figures and tables are available upon request.

### Topographical electrode power comparison

Cluster-based permutation analysis^98^ was used to identify any differences in relative or absolute power between FXS and controls. Overall alpha was set at .05 / 7 (adj. p.<.007) to account for multiple frequency band comparisons (effective alpha for each tail .025).

### Source model power comparison

Group-level statistical (t-statistic) cortical maps were generated by Monte-Carlo permutation (2000) after independent two-tailed t-tests (alpha set at .025 per tail) using the ‘ft_sourcestatistics’ function in FieldTrip^99^ and threshold at p<.05. The resulting p values were globally corrected by a false discovery rate (FDR) of 5% applied over the signals and frequency band dimensions^100^.

### Node, Region, and RSN Comparisons

Log-transformed power differences were evaluated with generalized linear mixed effect models via NLME library in R4.1 and confirmed with GLIMMIX procedure in SAS 9.4 (SAS Institute Inc., Cary, NC, USA) in which random effect was subject and independent variables varied based on model. Fixed effects included GROUP (FXS vs. control), SEX (male vs. female), or FREQUENCY BAND (delta, theta, alpha1, alpha2, beta, gamma1, and gamma2). When specified, NODE indicates the 68 cortical parcels of the DK atlas, REGION refers to the 14 node groups that represent cortical lobes, and RSN refers to the six functional grouping of nodes (DMN, VIS, DAN, SAN, AUD, Other). In REGION and RSN models, nodes were treated as replicates. To ensure optimal model fit, we examined various structures of intra-subject covariance and link functions. For each model, plots based on the studentized residuals were examined. See Appendix 1 for a visual DK atlas key and Appendix 2 for comprehensive classification table of cortical nodes.

### Cross frequency amplitude coupling

To examine potential dependence between low-frequency power and high-frequency activity, we calculated CFC as previously reporting^12^. CFC was calculated based upon the mean whole brain power of each low frequency band (theta, alpha1, and alpha2) compared to gamma1 power within each individual cortical node. The Spearman’s correlation coefficient for each low frequency and gamma1 was calculated using the time series of mean relative power across 2-s epochs. Fisher’s Z-transform was used to normalize group-wise comparisons.

### Correlation analysis

As a successive step, frequency bands of significance were linearly correlated with clinical and behavioral measures. Primary Analysis: Shapiro-Wilk’s normality test was performed on variables to assess suitability for either Spearman’s rank-order or Pearson’s correlation test. A priori hypotheses for high-frequency bands (beta, gamma1, and gamma2) and low-frequency bands (theta, alpha1, and alpha2) with clinical variables were assessed with correlation tests with p values adjusted by FDR for multiple test iterations and partial correlations were used to adjust all correlations for age.

## Data Availability

EEG data is available to the public as federally mandated at the National Database for Autism Research (NDAR). Analysis code is in preparation for upload into public repositories and is available on request.

https://nda.nih.gov/edit_collection.html?id=2373

## Acknowledgments

We thank the participants and families who participated in this study. We would also like to thank Nicole Friedman, Michael Hong, Danielle Chin, and Janna Guilfoyle, who assisted with the project. The present study was federally funded by the National Institutes of Health (NIH) Fragile X Centers (U54HD082008 and U54HD104461).

