## Supplemental Materials for "Neocortical Localization and Thalamocortical Modulation of Neuronal Hyperexcitability in Fragile X Syndrome"

**Figure S1. Group Contrasts of Topographical Power of Scalp EEG by Frequency Band**

Group-level t-maps (5% FDR-corrected) of cluster permutation statistical comparison between FXS (n=70) and TDC (n=71) of scalp (electrode-level) EEG power As findings across scalp EEG remain spatially ambiguous[1], the findings were primarily used to generate hypotheses and assess for frequency specific trends across groups. Warmer colors indicate significant FXS>TDC, cooler colors indicate significant FXS<TDC, and gray areas indicate non-significant group differences.

A. Relative Power (FXS-TDC)

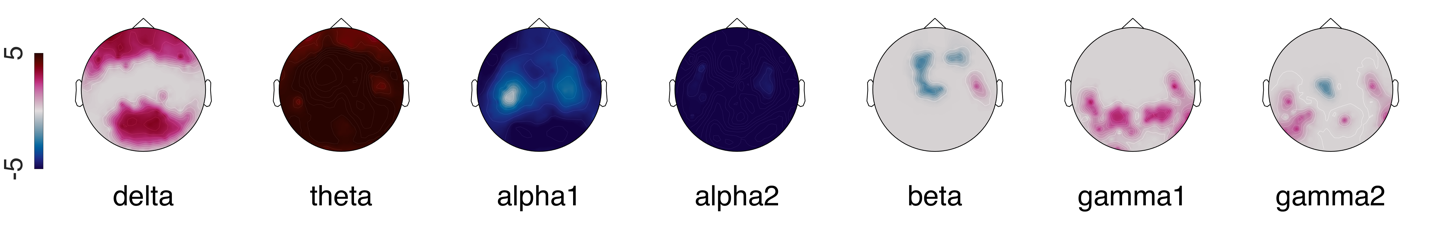

B. Absolute Power (FXS-TDC)

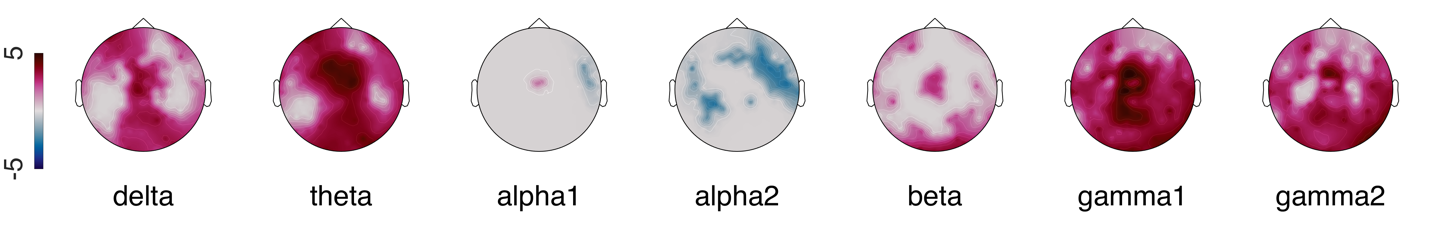
**Figure S2. Group Contrasts of Source-level Absolute Power by Frequency Band**

Group level t-maps depicting FXS – Control, vertex-by-vertex absolute power differences by frequency band superimposed on brain surface models. Warmer (FXS > Control) and cooler (FXS < Control) color scale represents significant t-values (non-significant values as gray).

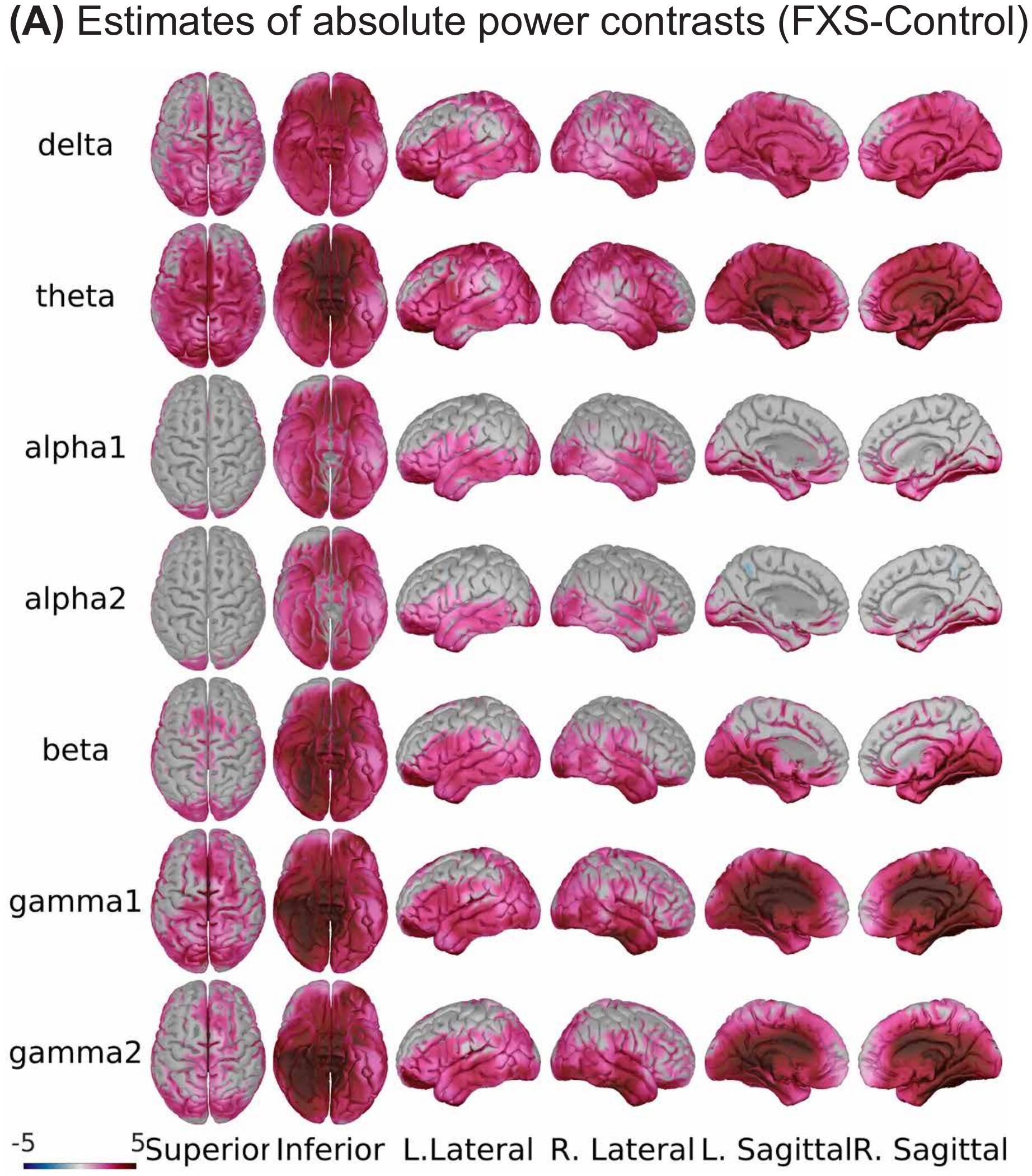

Supplemental Tables

Table S1: Additional Clinical Characteristics by Group

Comparison of detailed demographic and clinical features of EEG dataset by group. FSIQ, Full Scale IQ, NVIQ, Non-verbal intelligence scale; VIQ, verbal intelligence scale; SCQ, Social Communication Questionnaire; WJ-3, Woodcock III Tests of Cognitive Abilities; ABC, Aberrant Behavioral Checklist; ADAMS, Anxiety, Depression, and Mood Scale; t, t-statistic following independent Student t-tests; p, unadjusted significance; adj. p, p-value following Bonferroni correction.

|  | **Control(M)** | **FXS(M)** | **Control(F)** | **FXS(F)** | **p.overall** |
| --- | --- | --- | --- | --- | --- |
|  | ***N=41*** | ***N=38*** | ***N=30*** | ***N=32*** |  |
| Age (Years) | 22.2 (9.81) | 22.7 (9.95) | 22.0 (12.0) | 17.9 (9.61) | 0.200 |
| FSIQ | 106 (10.1) | 34.5 (22.8) | 99.9 (6.78) | 65.5 (29.2) | <0.001 |
| VIQ | 108 (13.3) | 46.9 (24.2) | 97.0 (9.38) | 71.7 (27.9) | <0.001 |
| NVIQ | 104 (12.2) | 22.1 (28.4) | 103 (8.65) | 59.2 (32.9) | <0.001 |
| SCQ | 1.73 (1.93) | 17.1 (6.49) | 2.65 (2.52) | 9.36 (7.00) | <0.001 |
| ABC-Irritability | 0.47 (0.97) | 11.5 (9.30) | 1.12 (2.66) | 7.21 (7.87) | <0.001 |
| ABC-Hyperactivity | 0.90 (1.21) | 18.3 (11.3) | 1.25 (4.23) | 8.21 (8.02) | <0.001 |
| ABC-Abnormal Speech | 0.10 (0.31) | 5.88 (3.29) | 0.19 (0.75) | 2.50 (2.74) | <0.001 |
| ABC-Lethargy | 1.30 (3.23) | 8.39 (6.76) | 0.81 (2.48) | 7.50 (8.92) | <0.001 |
| ABC-Stereotypy | 0.03 (0.18) | 6.30 (5.56) | 0.12 (0.50) | 1.86 (3.82) | <0.001 |
| ADAMS-OCD | 0.23 (0.68) | 3.14 (2.66) | 0.56 (1.54) | 1.29 (1.84) | <0.001 |
| ADAMS-Anxiety | 2.03 (2.22) | 7.83 (5.01) | 2.28 (3.12) | 6.37 (5.07) | <0.001 |
| WJ-III | 95.3 (13.4) | 65.5 (15.8) | 91.3 (8.02) | 70.9 (15.4) | <0.001 |

**Table S2: Preprocessing.** Statistical comparison of preprocessing characteristics demonstrating no significant group differences in key artifact cleaning routines across final datasets.

|  | **FXS** | **Control** | **p** |
| --- | --- | --- | --- |
|  | ***N=70*** | ***N=71*** |  |
| Remaining Trials | 125 (24.6) | 131 (18.1) | 0.117 |
| Total Duration(s) | 315 (29.0) | 311 (25.9) | 0.437 |
| Clean Duration(s) | 250 (49.2) | 262 (36.1) | 0.115 |
| Bad Channels | 6.27 (3.31) | 5.93 (3.71) | 0.564 |
| Artifact Components | 11.3 (1.71) | 10.8 (2.58) | 0.117 |

**Table S3: Summary of Source-estimated Peak Alpha Frequency (PAF) by Cortical Node.** A linear mixed effect model was conducted to examine the effect of group, sex, and cortical node location on source estimated PAF (subject was a random effect). A significant interaction effect between group and node (group x node; F_67,9378_=1.44, p=0.01) was present, but no effect of sex. Each row summarizes 5% FDR corrected significant pairwise comparisons (FXS-Control) of PAF estimates (Hz). Atlas key in Appendix 1. Abbreviations: FXS, Fragile X Syndrome; Cont., control; SE, standard error; DF, degrees of freedom; FDR, false discovery rate.

| Node | Region | FXS | TDC | FXS-TDC | SE | DF | F | 5% FDR |
| --- | --- | --- | --- | --- | --- | --- | --- | --- |
| Banks of Sup. Temp. Sulcus R | RT | 8.24±.22 | 9.19±.22 | -.95 | .30 | 137 | -3.1 | 0.011 |
| Caudal Anterior Cingulate L | LL | 7.96±.22 | 8.72±.22 | -.76 | .31 | 137 | -2.5 | 0.037 |
| Caudal Anterior Cingulate R | RL | 7.74±.22 | 8.61±.22 | -.87 | .30 | 137 | -2.8 | 0.017 |
| Caudal Middle Frontal L | LF | 8.26±.22 | 9.29±.22 | -1.03 | .31 | 137 | -3.4 | 0.008 |
| Caudal Middle Frontal R | RF | 8.39±.22 | 9.21±.22 | -.82 | .30 | 137 | -2.7 | 0.024 |
| Cuneus L | LO | 7.86±.22 | 8.79±.22 | -.93 | .30 | 137 | -3.1 | 0.011 |
| Cuneus R | RO | 8.45±.22 | 9.46±.22 | -1.01 | .30 | 137 | -3.3 | 0.009 |
| Entorhinal L | LT | 7.61±.22 | 8.34±.22 | -.73 | .30 | 137 | -2.4 | 0.045 |
| Fusiform R | RT | 8.54±.22 | 9.28±.22 | -.74 | .30 | 137 | -2.4 | 0.041 |
| Inferior Parietal L | LP | 8.30±.22 | 9.26±.22 | -.97 | .30 | 137 | -3.2 | 0.011 |
| Insula L | LT | 8.29±.22 | 9.19±.22 | -.91 | .30 | 137 | -3.0 | 0.012 |
| Isthmus Cingulate L | LL | 8.40±.22 | 9.33±.22 | -.93 | .30 | 137 | -3.0 | 0.011 |
| Isthmus Cingulate R | RL | 8.27±.22 | 9.26±.22 | -.99 | .30 | 137 | -3.3 | 0.010 |
| Lateral Occipital L | LO | 8.22±.22 | 9.13±.22 | -.92 | .30 | 137 | -3.0 | 0.011 |
| Paracentral L | LC | 8.88±.22 | 9.60±.22 | -.72 | .30 | 137 | -2.4 | 0.045 |
| Paracentral R | RC | 8.68±.22 | 9.46±.22 | -.78 | .30 | 137 | -2.5 | 0.034 |
| Pars Opercularis L | LF | 7.79±.22 | 8.51±.22 | -.72 | .30 | 137 | -2.4 | 0.045 |
| Pars Triangularis R | RF | 7.68±.22 | 8.79±.22 | -1.12 | .30 | 137 | -3.7 | 0.004 |
| Postcentral L | LC | 8.25±.22 | 9.48±.22 | -1.23 | .30 | 137 | -4.0 | 0.002 |
| Postcentral R | RC | 8.31±.22 | 9.23±.22 | -.92 | .30 | 137 | -3.0 | 0.011 |
| Posteriorcingulate L | LL | 8.23±.22 | 9.17±.22 | -.94 | .31 | 137 | -3.1 | 0.011 |
| Posteriorcingulate R | RL | 8.00±.22 | 8.95±.22 | -.95 | .31 | 137 | -3.1 | 0.011 |
| Precentral L | LC | 8.19±.22 | 9.62±.22 | -1.43 | .30 | 137 | -4.7 | <0.001 |
| Precentral R | RC | 8.51±.22 | 9.26±.22 | -.75 | .31 | 137 | -2.4 | 0.041 |
| Precuneus L | LP | 8.07±.22 | 9.34±.22 | -1.27 | .30 | 137 | -4.2 | 0.001 |
| Precuneus R | RP | 8.31±.22 | 9.10±.22 | -.79 | .30 | 137 | -2.6 | 0.033 |
| Superior Parietal L | LP | 8.11±.22 | 9.26±.22 | -1.15 | .30 | 137 | -3.8 | 0.003 |
| Superior Parietal R | RP | 7.85±.22 | 9.21±.22 | -1.36 | .30 | 137 | -4.5 | <0.001 |
| Supramarginal L | LP | 8.72±.22 | 9.82±.22 | -1.10 | .30 | 137 | -3.6 | 0.004 |
| Supramarginal R | RP | 8.34±.22 | 9.30±.22 | -.95 | .30 | 137 | -3.1 | 0.011 |

**Table S4: Pairwise group differences of gamma1 cross-frequency coupling by cortical node.**

| Pairwise (FXS-Control) gamma1 power-power CFC comparisons by cortical node | | | | | | |
| --- | --- | --- | --- | --- | --- | --- |
|  | Theta | | Alpha1 | | Alpha2 | |
| Node | L | R | L | R | L | R |
| Banks of Sup. Temp. Sulcus | -.08* | -.12*** | .06* | .06* | .10** | .06* |
| Caudal Anterior Cingulate | -.11** | -.11** | .02 | .02 | .08* | .08* |
| Caudal Middle Frontal | -.11** | -.11*** | .03 | .03 | .07* | .05 |
| Cuneus | -.05* | -.09* | .06* | .08* | .09* | .12*** |
| Entorhinal | -.08* | -.05 | .06* | .08* | .08* | .10** |
| Frontal Pole | .02 | .01 | .05* | .06* | .06* | .06* |
| Fusiform | -.06* | -.07* | .08* | .09* | .09* | .12*** |
| Inferior Parietal | -.08* | -.11** | .04 | .07* | .10** | .08* |
| Inferior Temporal | -.08* | -.07* | .04 | .09* | .07* | .10** |
| Insula | -.08* | -.05* | .04 | .03 | .05 | .07* |
| Isthmus Cingulate | -.12*** | -.13*** | .10** | .11** | .15*** | .12*** |
| Lateral Occipital | -.03 | -.05* | .08* | .07* | .09* | .09* |
| Lateral Orbitofrontal | -.01 | -.04 | .11** | .06* | .10** | .09** |
| Lingual | -.03 | -.04 | .12*** | .09* | .08* | .09** |
| Medial Orbitofrontal | -.01 | -.05 | .08* | .06* | .08* | .08* |
| Middle Temporal | -.06* | -.06* | .07* | .07* | .07* | .10** |
| Paracentral | -.12*** | -.12*** | .04 | .03 | .12*** | .11*** |
| Parahippocampal | -.08* | -.09* | .09** | .09** | .09** | .11** |
| Pars Opercularis | -.06* | -.03 | .02 | .05 | .06* | .05* |
| Pars Orbitalis | -.02 | .03 | .07* | .03 | .08* | .05 |
| Pars Triangularis | -.05* | -.06* | .04 | .01 | .07* | .06* |
| Pericalcarine | -.03 | -.04 | .11** | .09** | .08* | .07* |
| Postcentral | -.12*** | -.10** | .06* | .05 | .07* | .05* |
| Posteriorcingulate | -.13*** | -.13*** | .04 | .05 | .10** | .09* |
| Precentral | -.11*** | -.13*** | .06* | .05 | .06* | .08* |
| Precuneus | -.12*** | -.14*** | .09* | .06* | .10** | .11*** |
| Rostral Anterior Cingulate | -.03 | -.03 | .07* | .06* | .06* | .07* |
| Rostral Middle Frontal | -.03 | -.04 | .06* | .02 | .06* | .05* |
| Superior Frontal | -.09** | -.08* | .01 | -.02 | .07* | .07* |
| Superior Parietal | -.13*** | -.13*** | .09* | .09* | .07* | .11** |
| Superior Temporal | -.07* | -.06* | .05 | .06* | .06* | .09* |
| Supramarginal | -.11** | -.10** | .05* | .04 | .08* | .06* |
| Temporal Pole | -.04 | -.05 | .05 | .08* | .05* | .10** |
| Transverse Temporal | -.08* | -.08* | .07* | .05* | .07* | .09** |

**Table S5: Pairwise group differences of gamma1 cross-frequency coupling by resting state network (RSN).**

| RSN | Lower Band | Estimate | Statistic | DF | 5% FDR p | Sig. |
| --- | --- | --- | --- | --- | --- | --- |
| DMN | theta | -.08±.01 | -6.26 | 139 | 1.6e-08 | *** |
|  | alpha1 | .07±.01 | 5.62 | 139 | 2.3e-07 | *** |
|  | alpha2 | .09±.01 | 7.34 | 139 | 2.9e-10 | *** |
| DAN | theta | -.05±.01 | -3.91 | 139 | 2.0e-04 | *** |
|  | alpha1 | .05±.01 | 3.48 | 139 | 8.1e-04 | *** |
|  | alpha2 | .07±.01 | 5.41 | 139 | 5.4e-07 | *** |
| SAN | theta | -.08±.01 | -5.64 | 139 | 2.3e-07 | *** |
|  | alpha1 | .04±.01 | 2.65 | 139 | 9.0e-03 | ** |
|  | alpha2 | .06±.01 | 4.37 | 139 | 3.9e-05 | *** |
| AUD | theta | -.07±.02 | -3.41 | 139 | 9.6e-04 | *** |
|  | alpha1 | .05±.02 | 2.82 | 139 | 5.8e-03 | ** |
|  | alpha2 | .08±.02 | 3.96 | 139 | 1.8e-04 | *** |
| VIS | theta | -.05±.01 | -3.79 | 139 | 2.9e-04 | *** |
|  | alpha1 | .08±.01 | 5.86 | 139 | 9.6e-08 | *** |
|  | alpha2 | .09±.01 | 6.80 | 139 | 2.6e-09 | *** |
| other | theta | -.08±.01 | -6.49 | 139 | 8.5e-09 | *** |
|  | alpha1 | .06±.01 | 4.65 | 139 | 1.4e-05 | *** |
|  | alpha2 | .08±.01 | 6.38 | 139 | 1.1e-08 | *** |

Table S6: Age-corrected Clinical Correlations of Spectral Power by Cortical Region

Abbreviations: See Appendix 2 for region atlas key; ABC, Aberrant Behavior Checklist; ADAMS, Anxiety, Depression and Mood Scale; SCQ, Social Communication Questionnaire; VIQ, verbal intelligence quotient.

All FXS Participants

|  | | Spearman's ρ | | | | Age-Corrected | |
| --- | --- | --- | --- | --- | --- | --- | --- |
| Measure | Frequency | Region | n | rho | adj.p | rho | adj.p |
| ABC-Abnormal Speech | alpha1 | LPF | 61 | -.44 | 0.01 | -.48 | <0.01 |
|  |  | RPF | 61 | -.40 | 0.03 | -.44 | 0.02 |
|  | alpha2 | LPF | 61 | -.35 | 0.05 | -.38 | 0.04 |
| ABC-Hyperactivity | alpha1 |  | 61 | -.42 | 0.02 | -.40 | 0.03 |
|  |  | LT | 61 | -.46 | 0.01 | -.44 | 0.02 |
|  |  | RPF | 61 | -.44 | 0.01 | -.42 | 0.02 |
|  |  | RT | 61 | -.49 | <0.01 | -.48 | <0.01 |
| ABC-Stereotypy | theta | LPF | 61 | -.40 | 0.03 | -.40 | 0.03 |
|  |  | RPF | 61 | -.39 | 0.03 | -.39 | 0.04 |
|  | alpha1 | LO | 61 | -.30 | 0.1 | -.39 | 0.04 |
|  |  | LPF | 61 | -.47 | <0.01 | -.49 | <0.01 |
|  |  | LT | 61 | -.32 | 0.08 | -.39 | 0.04 |
|  |  | RO | 61 | -.33 | 0.07 | -.41 | 0.03 |
|  |  | RPF | 61 | -.48 | <0.01 | -.50 | <0.01 |
|  |  | RT | 61 | -.38 | 0.04 | -.45 | 0.01 |
|  | alpha2 | LPF | 61 | -.44 | 0.01 | -.46 | 0.01 |
|  |  | RO | 61 | -.32 | 0.08 | -.39 | 0.04 |
|  |  | RPF | 61 | -.39 | 0.03 | -.41 | 0.03 |
| ADAMS-OCD | alpha1 | LO | 64 | -.39 | 0.03 | -.42 | 0.02 |
|  |  | LP | 64 | -.36 | 0.04 | -.39 | 0.03 |
|  |  | LPF | 64 | -.39 | 0.03 | -.39 | 0.03 |
|  |  | LT | 64 | -.44 | 0.01 | -.47 | <0.01 |
|  |  | RO | 64 | -.36 | 0.05 | -.38 | 0.04 |
|  |  | RP | 64 | -.36 | 0.04 | -.39 | 0.03 |
|  |  | RPF | 64 | -.43 | 0.01 | -.44 | 0.02 |
|  |  | RT | 64 | -.45 | <0.01 | -.48 | <0.01 |
|  | alpha2 | LC | 64 | -.39 | 0.03 | -.39 | 0.03 |
|  |  | LL | 64 | -.49 | <0.01 | -.49 | <0.01 |
|  |  | LP | 64 | -.40 | 0.03 | -.42 | 0.02 |
|  |  | LPF | 64 | -.45 | <0.01 | -.45 | 0.01 |
|  |  | LT | 64 | -.40 | 0.02 | -.42 | 0.02 |
|  |  | RC | 64 | -.40 | 0.02 | -.40 | 0.03 |
|  |  | RL | 64 | -.48 | <0.01 | -.48 | <0.01 |
|  |  | RP | 64 | -.42 | 0.02 | -.43 | 0.02 |
|  |  | RPF | 64 | -.44 | 0.01 | -.44 | 0.01 |
|  |  | RT | 64 | -.42 | 0.02 | -.44 | 0.02 |
| SCQ | alpha1 | LPF | 62 | -.46 | <0.01 | -.48 | <0.01 |
|  |  | LT | 62 | -.35 | 0.06 | -.41 | 0.03 |
|  |  | RPF | 62 | -.46 | <0.01 | -.49 | <0.01 |
|  |  | RT | 62 | -.33 | 0.07 | -.39 | 0.04 |
|  | alpha2 | LF | 62 | -.38 | 0.03 | -.39 | 0.04 |
|  |  | LL | 62 | -.40 | 0.03 | -.42 | 0.02 |
|  |  | LPF | 62 | -.51 | <0.01 | -.52 | <0.01 |
|  |  | RC | 62 | -.36 | 0.05 | -.37 | 0.05 |
|  |  | RL | 62 | -.40 | 0.03 | -.43 | 0.02 |
|  |  | RPF | 62 | -.47 | <0.01 | -.49 | <0.01 |
| VIQ | alpha1 | LPF | 64 | .35 | 0.05 | .40 | 0.03 |

Table S7: Age-corrected Clinical Correlations of Spectral Power by Cortical Region

Abbreviations: See Appendix 2 for region atlas key; ABC, Aberrant Behavior Checklist; ADAMS, Anxiety, Depression and Mood Scale; SCQ, Social Communication Questionnaire; WJ-3, Woodcock-Johnson III Tests of Cognitive Abilities, Auditory Attention subscale; NVIQ, non-verbal intelligence quotient; VIQ, verbal intelligence quotient.

Full mutation, non-mosaic males with FXS only

|  | | Spearman's ρ | | | | Age-Corrected | |
| --- | --- | --- | --- | --- | --- | --- | --- |
| Measure | Frequency | Region | n | rho | p | rho | p |
| ABC-Abnormal Speech | theta | LPF | 24 | -.48 | 0.02 | -.50 | 0.01 |
|  |  | RPF | 24 | -.48 | 0.02 | -.49 | 0.02 |
|  | alpha1 | LPF | 24 | -.50 | 0.01 | -.49 | 0.02 |
|  |  | RPF | 24 | -.44 | 0.03 | -.42 | 0.05 |
|  | gamma1 | LPF | 24 | .36 | 0.08 | .42 | 0.05 |
|  |  | RF | 24 | .46 | 0.03 | .50 | 0.02 |
|  |  | RL | 24 | .44 | 0.03 | .46 | 0.03 |
|  |  | RP | 24 | .46 | 0.02 | .49 | 0.02 |
|  |  | RPF | 24 | .46 | 0.02 | .50 | 0.02 |
|  |  | RT | 24 | .39 | 0.06 | .47 | 0.02 |
|  | gamma2 | LL | 24 | .43 | 0.04 | .48 | 0.02 |
|  |  | LPF | 24 | .38 | 0.07 | .45 | 0.03 |
|  |  | RC | 24 | .39 | 0.06 | .45 | 0.03 |
|  |  | RL | 24 | .44 | 0.03 | .47 | 0.02 |
|  |  | RP | 24 | .57 | <0.01 | .60 | <0.01 |
|  |  | RPF | 24 | .54 | <0.01 | .58 | <0.01 |
|  |  | RT | 24 | .54 | <0.01 | .59 | <0.01 |
| ABC-Hyperactivity | gamma1 | LF | 24 | .08 | 0.72 | .43 | 0.04 |
|  |  | RP | 24 | .26 | 0.23 | .42 | 0.04 |
| ABC-Irritability |  |  | 24 | .34 | 0.1 | .42 | 0.04 |
| ABC-Lethargy | theta | LPF | 24 | -.46 | 0.02 | -.48 | 0.02 |
|  |  | RPF | 24 | -.49 | 0.02 | -.49 | 0.02 |
|  | alpha1 | LPF | 24 | -.46 | 0.02 | -.45 | 0.03 |
|  |  | RPF | 24 | -.53 | <0.01 | -.51 | 0.01 |
|  | alpha2 | LPF | 24 | -.46 | 0.02 | -.44 | 0.04 |
|  |  | RPF | 24 | -.51 | 0.01 | -.49 | 0.02 |
|  | gamma1 | LF | 24 | .33 | 0.11 | .43 | 0.04 |
| ABC-Stereotypy | theta | LC | 24 | -.33 | 0.12 | -.41 | 0.05 |
|  |  | LF | 24 | -.40 | 0.05 | -.49 | 0.02 |
|  |  | LPF | 24 | -.53 | <0.01 | -.60 | <0.01 |
|  |  | RC | 24 | -.33 | 0.11 | -.45 | 0.03 |
|  |  | RP | 24 | -.30 | 0.15 | -.43 | 0.04 |
|  |  | RPF | 24 | -.55 | <0.01 | -.59 | <0.01 |
|  |  | RT | 24 | -.40 | 0.06 | -.43 | 0.04 |
|  | alpha1 | LPF | 24 | -.64 | <0.01 | -.66 | <0.01 |
|  |  | RO | 24 | -.51 | 0.01 | -.43 | 0.04 |
|  |  | RPF | 24 | -.62 | <0.01 | -.62 | <0.01 |
|  | alpha2 | LPF | 24 | -.56 | <0.01 | -.55 | <0.01 |
|  |  | RPF | 24 | -.51 | 0.01 | -.48 | 0.02 |
| ADAMS-Anxiety | alpha1 |  | 27 | -.48 | 0.01 | -.46 | 0.02 |
|  |  | RT | 27 | -.45 | 0.02 | -.39 | 0.05 |
|  | alpha2 | LO | 27 | -.49 | <0.01 | -.43 | 0.03 |
|  |  | LPF | 27 | -.49 | <0.01 | -.47 | 0.02 |
|  |  | LT | 27 | -.58 | <0.01 | -.54 | <0.01 |
|  |  | RO | 27 | -.60 | <0.01 | -.56 | <0.01 |
|  |  | RPF | 27 | -.48 | 0.01 | -.46 | 0.02 |
|  |  | RT | 27 | -.48 | 0.01 | -.42 | 0.03 |
| ADAMS-OCD | alpha1 | LO | 27 | -.53 | <0.01 | -.45 | 0.02 |
|  |  | LT | 27 | -.56 | <0.01 | -.48 | 0.01 |
|  |  | RO | 27 | -.47 | 0.01 | -.40 | 0.04 |
| NVIQ | alpha2 | LT | 22 | -.50 | 0.02 | -.46 | 0.04 |
|  | gamma1 | LP | 22 | -.45 | 0.04 | -.44 | 0.04 |
|  |  | LT | 22 | -.51 | 0.01 | -.48 | 0.03 |
|  |  | RO | 22 | -.62 | <0.01 | -.60 | <0.01 |
|  |  | RT | 22 | -.52 | 0.01 | -.49 | 0.02 |
|  | gamma2 | LP | 22 | -.47 | 0.03 | -.46 | 0.04 |
|  |  | LT | 22 | -.53 | 0.01 | -.51 | 0.02 |
|  |  | RO | 22 | -.59 | <0.01 | -.57 | <0.01 |
|  |  | RT | 22 | -.55 | <0.01 | -.54 | 0.01 |
| SCQ | alpha1 | RPF | 24 | -.44 | 0.03 | -.43 | 0.04 |
|  | alpha2 |  | 24 | -.49 | 0.01 | -.46 | 0.03 |
| VIQ |  | LT | 22 | -.30 | 0.17 | -.57 | <0.01 |
|  |  | RT | 22 | -.25 | 0.26 | -.46 | 0.04 |
| WJ-III | theta | LO | 23 | .32 | 0.14 | .45 | 0.04 |
|  |  | LP | 23 | .42 | 0.05 | .46 | 0.03 |
|  |  | RO | 23 | .31 | 0.15 | .45 | 0.04 |
|  |  | RP | 23 | .38 | 0.07 | .52 | 0.01 |
|  | alpha1 | LO | 23 | .60 | <0.01 | .52 | 0.01 |
|  | gamma1 |  | 23 | -.17 | 0.44 | -.43 | 0.05 |
|  |  | RP | 23 | -.41 | 0.05 | -.49 | 0.02 |
|  | gamma2 | LO | 23 | -.22 | 0.31 | -.48 | 0.02 |
|  |  | RO | 23 | -.29 | 0.18 | -.43 | 0.04 |
|  |  | RP | 23 | -.41 | 0.05 | -.49 | 0.02 |

Table S8: Age-corrected Clinical Correlations of Spectral Power by Resting State Network (RSN)

Abbreviations: ABC, Aberrant Behavior Checklist; ADAMS, Anxiety, Depression and Mood Scale; SCQ, Social Communication Questionnaire; WJ-3, Woodcock-Johnson III Tests of Cognitive Abilities, Auditory Attention subscale; Map, RSN or region label; DMN, default mode network; DAN, dorsal attention network; SAN, salient affective network; VIS, visual attention network; AUD, auditory network.

All FXS Participants

|  | | Spearman's ρ | | | | Age-Corrected | |
| --- | --- | --- | --- | --- | --- | --- | --- |
| Measure | Frequency | RSN | n | rho | adj.p | rho | adj.p |
| ABC-Abnormal Speech | gamma2 | SAN | 61 | .38 | 0.03 | .36 | 0.05 |
| ABC-Hyperactivity | alpha1 | AUD | 61 | -.46 | 0.01 | -.44 | 0.02 |
|  |  | DAN | 61 | -.42 | 0.02 | -.40 | 0.03 |
| ABC-Stereotypy | theta | AUD | 61 | -.38 | 0.03 | -.38 | 0.04 |
|  | alpha1 |  | 61 | -.36 | 0.03 | -.40 | 0.03 |
|  |  | DAN | 61 | -.36 | 0.03 | -.39 | 0.03 |
|  |  | VIS | 61 | -.36 | 0.03 | -.44 | 0.02 |
|  | alpha2 | DAN | 61 | -.34 | 0.04 | -.37 | 0.04 |
|  |  | DMN | 61 | -.34 | 0.04 | -.39 | 0.03 |
|  |  | VIS | 61 | -.30 | 0.08 | -.38 | 0.04 |
| ADAMS-OCD | alpha1 | AUD | 64 | -.45 | 0.01 | -.46 | 0.02 |
|  |  | DAN | 64 | -.38 | 0.03 | -.38 | 0.03 |
|  |  | DMN | 64 | -.39 | 0.02 | -.41 | 0.03 |
|  |  | VIS | 64 | -.39 | 0.02 | -.42 | 0.02 |
|  | alpha2 | AUD | 64 | -.41 | 0.02 | -.42 | 0.02 |
|  |  | DAN | 64 | -.38 | 0.03 | -.38 | 0.03 |
|  |  | DMN | 64 | -.50 | <0.01 | -.52 | <0.01 |
|  |  | SAN | 64 | -.41 | 0.02 | -.42 | 0.02 |
|  |  | VIS | 64 | -.33 | 0.04 | -.36 | 0.05 |
| SCQ | alpha1 | AUD | 62 | -.32 | 0.05 | -.36 | 0.05 |
|  |  | DAN | 62 | -.37 | 0.03 | -.40 | 0.03 |
|  |  | DMN | 62 | -.34 | 0.04 | -.40 | 0.03 |
|  |  | SAN | 62 | -.33 | 0.05 | -.36 | 0.05 |
|  | alpha2 | DAN | 62 | -.36 | 0.03 | -.39 | 0.03 |
|  |  | DMN | 62 | -.39 | 0.03 | -.44 | 0.02 |
|  |  | SAN | 62 | -.39 | 0.02 | -.41 | 0.03 |
| WJ-III | alpha1 |  | 58 | .37 | 0.03 | .39 | 0.03 |

Table S9: Age-corrected Clinical Correlations of Spectral Power by Resting State Network

Abbreviations: ABC, Aberrant Behavior Checklist; ADAMS, Anxiety, Depression and Mood Scale; WJ-3, Woodcock-Johnson III Tests of Cognitive Abilities, Auditory Attention subscale; NVIQ, non-verbal intelligence quotient; Map, RSN or region label; DMN, default mode network; DAN, dorsal attention network; SAN, salient affective network; VIS, visual attention network; AUD, auditory network.

Full mutation, non-mosaic males with FXS only

|  | | Spearman's ρ | | | | Age-Corrected | |
| --- | --- | --- | --- | --- | --- | --- | --- |
| Measure | Frequency | Rsn | n | rho | p | rho | p |
| ABC-Abnormal Speech | alpha2 | SAN | 24 | .37 | 0.08 | .51 | 0.01 |
|  | gamma1 | AUD | 24 | .34 | 0.1 | .43 | 0.04 |
|  |  | DAN | 24 | .42 | 0.04 | .51 | 0.01 |
|  |  | DMN | 24 | .39 | 0.06 | .43 | 0.04 |
|  |  | SAN | 24 | .40 | 0.05 | .45 | 0.03 |
|  | gamma2 | AUD | 24 | .44 | 0.03 | .49 | 0.02 |
|  |  | DAN | 24 | .46 | 0.02 | .53 | <0.01 |
|  |  | DMN | 24 | .46 | 0.02 | .51 | 0.01 |
| ABC-Lethargy | theta | AUD | 24 | -.41 | 0.05 | -.45 | 0.03 |
| ABC-Stereotypy |  |  | 24 | -.37 | 0.08 | -.47 | 0.02 |
|  |  | DAN | 24 | -.38 | 0.07 | -.43 | 0.04 |
|  |  | DMN | 24 | -.46 | 0.02 | -.46 | 0.03 |
|  |  | SAN | 24 | -.39 | 0.06 | -.47 | 0.03 |
|  | alpha1 | DAN | 24 | -.47 | 0.02 | -.44 | 0.04 |
| ADAMS-Anxiety | alpha2 | VIS | 27 | -.55 | <0.01 | -.51 | <0.01 |
| ADAMS-OCD | alpha1 | AUD | 27 | -.49 | <0.01 | -.44 | 0.03 |
|  |  | VIS | 27 | -.51 | <0.01 | -.41 | 0.04 |
|  | alpha2 | DMN | 27 | -.50 | <0.01 | -.40 | 0.04 |
| NVIQ | gamma1 | AUD | 22 | -.49 | 0.02 | -.45 | 0.04 |
|  |  | DAN | 22 | -.48 | 0.02 | -.44 | 0.04 |
|  |  | DMN | 22 | -.47 | 0.03 | -.46 | 0.04 |
|  |  | VIS | 22 | -.63 | <0.01 | -.60 | <0.01 |
|  | gamma2 | AUD | 22 | -.54 | <0.01 | -.52 | 0.02 |
|  |  | DMN | 22 | -.45 | 0.03 | -.44 | 0.05 |
|  |  | VIS | 22 | -.53 | 0.01 | -.50 | 0.02 |
| WJ-III | gamma1 |  | 23 | -.23 | 0.3 | -.42 | 0.05 |
|  | gamma2 |  | 23 | -.25 | 0.25 | -.44 | 0.04 |

**Table S10: Age-corrected Clinical Correlations of Peak Alpha Frequency (PAF) by Cortical Region and Resting State Network (RSN).**

Abbreviations: ADAMS, Anxiety, Depression and Mood Scale; VIQ, verbal intelligence quotient; DMN, default mode network; SAN, salient affective network.

All FXS Participants (trending significance following FDR correction)

| Measure | RSN | Type | r | p | adj. p | n |
| --- | --- | --- | --- | --- | --- | --- |
| VIQ | SAN | PAF | .39 | 1.5e-03 | 0.081 | 64 |

**Full mutation, non-mosaic males with FXS only**

**A. Cortical Regions**

|  | Spearman's ρ | | | | Age-Corrected | |
| --- | --- | --- | --- | --- | --- | --- |
| Measure | Region | n | rho | p | rho | p |
| ABC-Hyperactivity | LC | 24 | .14 | 0.53 | .42 | 0.05 |
|  | LF | 24 | -.50 | 0.01 | -.48 | 0.02 |
|  | LO | 24 | -.54 | <0.01 | -.42 | 0.05 |
| ABC-Stereotypy |  | 24 | -.54 | <0.01 | -.45 | 0.03 |
|  | RPF | 24 | .54 | <0.01 | .54 | <0.01 |
| ADAMS-Anxiety | LO | 27 | -.57 | <0.01 | -.53 | <0.01 |
|  | LPF | 27 | -.48 | 0.01 | -.43 | 0.03 |
|  | LT | 27 | -.53 | <0.01 | -.53 | <0.01 |
|  | RP | 27 | -.42 | 0.03 | -.41 | 0.04 |
|  | RT | 27 | -.42 | 0.03 | -.46 | 0.02 |

**B. RSNs**

|  | Spearman's ρ | | | | Age-Corrected | |
| --- | --- | --- | --- | --- | --- | --- |
| Measure | RSN | n | rho | p | rho | p |
| ADAMS-Anxiety | DMN | 27 | -.44 | 0.02 | -.46 | 0.02 |

Table S11: Age-corrected Clinical Correlations of Power-Power Cross-frequency Coupling (CFC) by Cortical Region and Resting State Network (RSN).

Abbreviations: ADAMS, Anxiety, Depression and Mood Scale; NVIQ, non-verbal intelligence quotient; Map, RSN or region label; DMN, default mode network; DAN, dorsal attention network; SAN, salient affective network; VIS, visual attention network; AUD, auditory network; LO, left occipital; RO, right occipital.

All FXS Participants: No significant correlations following FDR correction.

**Full mutation, non-mosaic males with FXS only**

**A. Cortical Regions**

|  | | Spearman's ρ | | | | Age-Corrected | |
| --- | --- | --- | --- | --- | --- | --- | --- |
| Measure | Frequency | Region | n | rho | p | rho | p |
| ADAMS-Anxiety | alpha1 | RO | 27 | .43 | 0.02 | .44 | 0.03 |
| ADAMS-OCD | theta |  | 27 | .40 | 0.04 | .39 | 0.05 |
|  | alpha1 |  | 27 | .40 | 0.04 | .42 | 0.03 |
| NVIQ | theta | LF | 22 | .49 | 0.02 | .45 | 0.04 |
|  |  | LT | 22 | .55 | <0.01 | .51 | 0.02 |
|  | alpha1 | RO | 22 | .58 | <0.01 | .58 | <0.01 |
|  |  | RT | 22 | .60 | <0.01 | .58 | <0.01 |
|  | alpha2 | RO | 22 | .44 | 0.04 | .44 | 0.04 |
|  |  | RP | 22 | .42 | 0.05 | .44 | 0.04 |
|  |  | RT | 22 | .56 | <0.01 | .55 | <0.01 |
| VIQ | alpha1 | LT | 22 | -.53 | 0.01 | -.50 | 0.02 |
|  | alpha2 |  | 22 | -.56 | <0.01 | -.55 | <0.01 |
| WJ-III |  | RC | 23 | .42 | 0.05 | .47 | 0.03 |

**B. RSNs**

|  | | Spearman's ρ | | | | Age-Corrected | |
| --- | --- | --- | --- | --- | --- | --- | --- |
| Measure | Frequency | Rsn | n | rho | p | rho | p |
| ADAMS-Anxiety | alpha1 | VIS | 27 | .42 | 0.03 | .41 | 0.04 |
| ADAMS-OCD | alpha2 |  | 27 | .38 | 0.05 | .40 | 0.04 |
| NVIQ | theta | DAN | 22 | .47 | 0.03 | .44 | 0.05 |
|  |  | SAN | 22 | .52 | 0.01 | .49 | 0.03 |
|  | alpha1 | AUD | 22 | .48 | 0.02 | .46 | 0.04 |
|  |  | DAN | 22 | .54 | <0.01 | .53 | 0.01 |
|  |  | VIS | 22 | .45 | 0.03 | .44 | 0.04 |
|  | alpha2 | DAN | 22 | .48 | 0.02 | .46 | 0.04 |

**Appendix 1: Node-level parcellation key for Desikan-Killiany Cortical Atlas.**

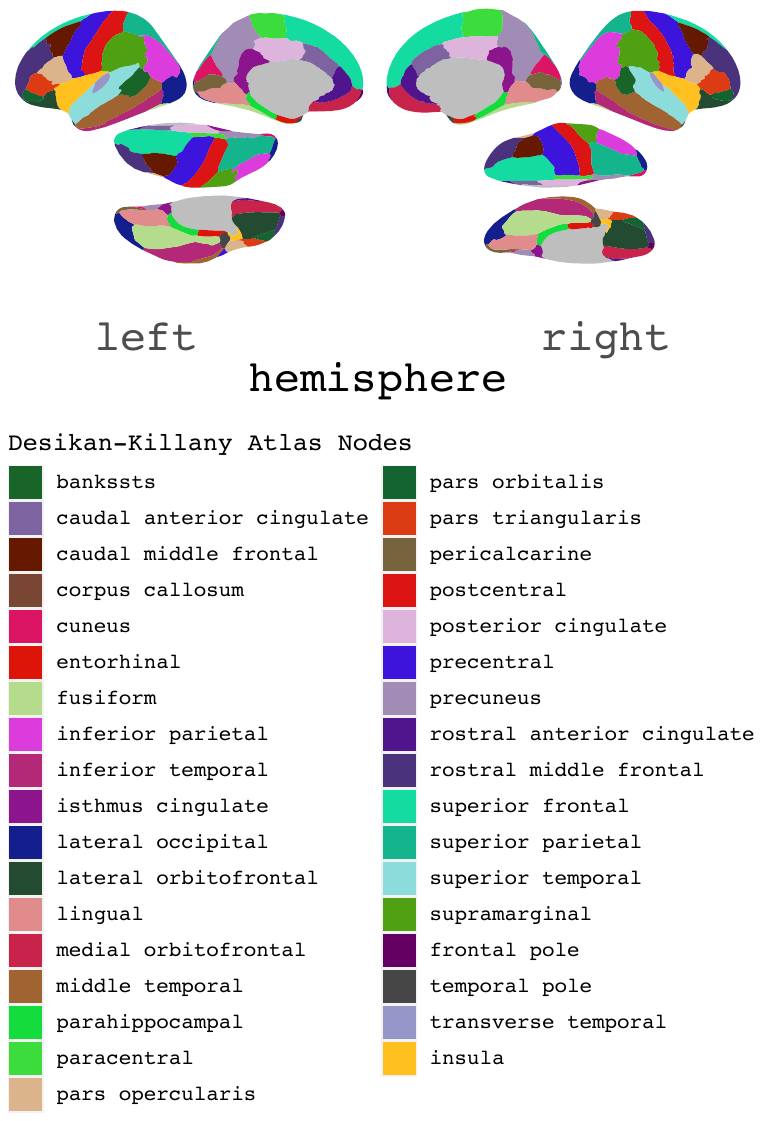

### Appendix 2: Assignment of cortical nodes to region and resting state network (RSN) groups as attributed by the Desikan-Killiany (DK) atlas. The MNI coordinates and number of vertices included in each node parcel are also displayed. Region and RSN represents an anatomical and functional grouping strategy for cortical nodes respectively.

### Abbreviations: RSN, resting state networks; DMN, default mode network; DAN, dorsal attention network; SAN, salience affective network; VIS, visual network; other, unassigned nodes. L, Left; R, right; F, frontal; L, lingula; O, occipital; P, parietal; PF, prefrontal; T, temporal.

|  |  |  |  | MNI Centroid | | |  |
| --- | --- | --- | --- | --- | --- | --- | --- |
| RSN | Node | Abbreviation | Cortex | x | y | z | Vert. |
| DMN | Caudal Anterior Cingulate R | cACC R | RL | 4 | 22 | 27 | 68 |
|  | Isthmus Cingulate L | iCC L | LL | -7 | -45 | 18 | 81 |
|  | Isthmus Cingulate R | iCC R | RL | 6 | -42 | 19 | 91 |
|  | Lateral Orbitofrontal L | LOF L | LPF | -25 | 30 | -19 | 221 |
|  | Lateral Orbitofrontal R | LOF R | RPF | 23 | 32 | -19 | 206 |
|  | Medial Orbitofrontal L | MOF L | LPF | -7 | 33 | -17 | 155 |
|  | Medial Orbitofrontal R | MOF R | RPF | 4 | 37 | -15 | 170 |
|  | Parahippocampal L | paraH L | LT | -26 | -29 | -21 | 68 |
|  | Parahippocampal R | paraH R | RT | 24 | -30 | -19 | 66 |
|  | Posterior Cingulate L | PCC L | LL | -5 | -15 | 38 | 85 |
|  | Posterior Cingulate R | PCC R | RL | 5 | -17 | 39 | 93 |
|  | Precuneus L | PCUN L | LP | -9 | -59 | 38 | 314 |
|  | Precuneus R | PCUN R | RP | 10 | -58 | 38 | 325 |
|  | Rostral Anterior Cingulate L | rACC L | LL | -5 | 39 | 1 | 78 |
|  | Rostral Anterior Cingulate R | rACC R | RL | 4 | 38 | 3 | 56 |
| SAN | Caudal Middle Frontal L | cMFG L | LF | -37 | 11 | 47 | 224 |
|  | Caudal Middle Frontal R | cMFG R | RF | 37 | 13 | 48 | 186 |
|  | Insula L | INS L | LT | -38 | -2 | 2 | 174 |
|  | Insula R | INS R | RT | 36 | 2 | -2 | 196 |
|  | Rostral Middle Frontal L | rMFG L | LF | -34 | 47 | 17 | 543 |
|  | Rostral Middle Frontal R | rMFG R | RF | 34 | 48 | 17 | 551 |
|  | Supramarginal L | SMAR L | LP | -57 | -38 | 34 | 305 |
|  | Supramarginal R | SMAR R | RP | 54 | -31 | 36 | 302 |
| DAN | Caudal Anterior Cingulate L | cACC L | LL | -5 | 21 | 26 | 48 |
|  | Inferior Temporal L | ITG L | LT | -53 | -36 | -22 | 307 |
|  | Inferior Temporal R | ITG R | RT | 51 | -32 | -25 | 316 |
|  | Middle Temporal L | MTG L | LT | -58 | -23 | -15 | 277 |
|  | Middle Temporal R | MTG R | RT | 58 | -22 | -15 | 324 |
|  | Pars Opercularis L | pOPER L | LF | -49 | 17 | 14 | 139 |
|  | Pars Opercularis R | pOPER R | RF | 49 | 17 | 14 | 118 |
|  | Pars Orbitalis L | pORB L | LPF | -44 | 39 | -14 | 72 |
|  | Pars Orbitalis R | pORB R | RPF | 43 | 42 | -15 | 68 |
|  | Pars Triangularis L | pTRI L | LF | -47 | 32 | 1 | 101 |
|  | Pars Triangularis R | pTRI R | RF | 48 | 34 | 2 | 148 |
| AUD | Superior Temporal L | STG L | LT | -55 | -12 | -4 | 290 |
|  | Superior Temporal R | STG R | RT | 54 | -6 | -7 | 257 |
| VIS | Cuneus L | CUN L | LO | -6 | -80 | 19 | 93 |
|  | Cuneus R | CUN R | RO | 8 | -78 | 20 | 99 |
|  | Fusiform L | FUS L | LT | -36 | -43 | -22 | 268 |
|  | Fusiform R | FUS R | RT | 35 | -41 | -23 | 255 |
|  | Lateral Occipital L | LOG L | LO | -31 | -89 | 0 | 371 |
|  | Lateral Occipital R | LOG R | RO | 35 | -85 | 2 | 367 |
|  | Lingual L | LING L | LO | -14 | -71 | -5 | 246 |
|  | Lingual R | LING R | RO | 13 | -67 | -4 | 227 |
| other | Banks of Sup. Temp. Sulcus L | BSTS L | LT | -53 | -45 | 8 | 76 |
|  | Banks of Sup. Temp. Sulcus R | BSTS R | RT | 54 | -41 | 10 | 70 |
|  | Entorhinal L | ENT L | LT | -26 | -5 | -33 | 30 |
|  | Entorhinal R | ENT R | RT | 23 | -6 | -35 | 32 |
|  | Frontal Pole L | FP L | LPF | -7 | 68 | -11 | 22 |
|  | Frontal Pole R | FP R | RPF | 7 | 68 | -15 | 30 |
|  | Inferior Parietal L | IPL L | LP | -42 | -71 | 32 | 351 |
|  | Inferior Parietal R | IPL R | RP | 46 | -63 | 32 | 421 |
|  | Paracentral L | paraC L | LC | -7 | -30 | 57 | 110 |
|  | Paracentral R | paraC R | RC | 7 | -27 | 57 | 128 |
|  | Pericalcarine L | periCAL L | LO | -11 | -82 | 6 | 109 |
|  | Pericalcarine R | periCAL R | RO | 12 | -80 | 7 | 110 |
|  | Postcentral L | postC L | LC | -46 | -22 | 45 | 333 |
|  | Postcentral R | postC R | RC | 44 | -20 | 46 | 307 |
|  | Precentral L | preC L | LC | -41 | -9 | 46 | 339 |
|  | Precentral R | preC R | RC | 40 | -7 | 46 | 353 |
|  | Superior Frontal L | sFG L | LF | -12 | 30 | 41 | 671 |
|  | Superior Frontal R | sFG R | RF | 12 | 32 | 41 | 603 |
|  | Superior Parietal L | SPL L | LP | -23 | -65 | 50 | 484 |
|  | Superior Parietal R | SPL R | RP | 24 | -65 | 51 | 464 |
|  | Temporal Pole L | TP L | LT | -28 | 14 | -38 | 38 |
|  | Temporal Pole R | TP R | RT | 27 | 16 | -36 | 38 |
|  | Transverse Temporal L | TT L | LT | -46 | -23 | 10 | 34 |
|  | Transverse Temporal R | TT R | RT | 46 | -17 | 9 | 23 |

1.	B, B., et al., *Spatial and Temporal Resolutions of EEG: Is It Really Black and White? A Scalp Current Density View.* International journal of psychophysiology : official journal of the International Organization of Psychophysiology, 2015. **97**(3).
